# Participatory Development of a Speech-Language Telerehabilitation Intervention combined with Home-based Transcranial Direct Current Stimulation for Primary Progressive Aphasia: a Qualitative Study

**DOI:** 10.1101/2024.12.10.24318315

**Authors:** Anna Uta Rysop, Roxana Schiwek, Tanja Grewe, Caterina Breitenstein, Ferdinand Binkofski, Mandy Roheger, Nina Unger, Agnes Flöel, Marcus Meinzer

## Abstract

**Purpose:** Intensive speech-language therapy (SLT) delivered as telerehabilitation in combination with transcranial direct current stimulation (tDCS) may be an effective treatment option for people with primary progressive aphasia (pwPPA). However, such combined treatment approaches are technically challenging and feasibility for pwPPA has not yet been established.

We aimed to involve stakeholders in the development of a novel approach combining remotely-supervised but independently self-administered home-based tDCS with an intensive aphasia telerehabilitation (naming and communicative-pragmatic therapy).

**Method:** Eight pwPPA and 16 caregivers participated in two semi-structured interviews to identify the needs, preferences, and perceived barriers and challenges with regard to SLT, telerehabilitation and tDCS, and the combination of all components. Based on the results, a step-by-step manual was developed and tested by means of home-based usability tests and follow-up interviews involving four dyads. Interview data were transcribed and analysed qualitatively and quantitatively. Competency checklists used in the usability tests were analysed quantitatively.

**Results:** PwPPA and their caregivers provided valuable insights into all aspects of the planned treatment programme. Overall, the findings suggest a high level of acceptance and perceived need for an intensive telerehabilitation SLT approach combined with tDCS. Using the developed step-by-step manual and training, pwPPA were able to independently perform more than half of the actions required for telerehabilitation but needed assistance with technically more demanding aspects of tDCS. To ensure feasibility, caregiver-assistance is needed to support pwPPA during technically challenging interventions.

**Conclusions:** This mixed-methods study identified needs and preferences of pwPPA and their caregivers with regard to speech-language telerehabilitation, as well as barriers and challenges regarding telerehabilitation and home-based tDCS. We demonstrate high acceptability and initial feasibility of such combined programmes. Our findings highlight the importance of stakeholder involvement in intervention development, which will inform future development and optimisation of technologically demanding intervention programmes.

## Introduction

Primary Progressive Aphasia (PPA) is a neurodegenerative disorder that primarily affects speech and language abilities, resulting in detrimental effects on communication and social participation (Mesulam, 1982, 2001). Typically, PPA can be classified into three variants, that differ in the underlying neuropathology, atrophy patterns and behavioural symptoms (Gorno-Tempini et al., 2011). These variants include semantic variant (svPPA), logopenic variant (lvPPA) and nonfluent agrammatic variant (nvPPA). Effective pharmacological treatment options are lacking, but behavioural approaches, such as speech-language therapy (SLT) have been suggested to ameliorate language impairment (Jokel et al., 2014; Volkmer et al., 2020; Wauters et al., 2023). Lexical naming interventions that focus on both semantic and phonological cues (Jokel et al., 2014), and script training aimed at improving communication abilities (Henry et al., 2019) have shown considerable and lasting beneficial effects on trained items (Wauters et al., 2023). In post-stroke aphasia, there is growing evidence that SLT is most effective when delivered at a moderate-to-high frequency and intensity (Brady et al., 2022; Pierce et al., 2024; Rose et al., 2022) and there is preliminary evidence that the same principle applies to PPA (Cadório et al., 2017; Wauters et al., 2023). However, intensive treatment schedules draw heavily on health care system resources and access to treatment may be difficult for patients with limited mobility and those living in rural areas.

While SLT is traditionally administered in person, telerehabilitation technology has advanced at a fast pace over the last years, making it a viable alternative option for treatment delivery (Cetinkaya et al., 2023; Suárez-González et al., 2024). Telerehabilitation refers to remotely-delivered treatment that can be either synchronous (i.e., internet-based via videoconferencing) or asynchronous (i.e., application-based). Initial evidence indicates that synchronous SLT is feasible and acceptable for people with PPA (pwPPA; Mooney et al., 2018; Rogalski et al., 2016; Rogalski et al., 2024) and is not inferior to in-person SLT in pwPPA (Dial et al., 2019; Meyer et al., 2016) and people with stroke-based aphasia (Jewell et al., 2024). However, telerehabilitation is technically demanding and requires a certain level of digital literacy.

Treatment success may be further enhanced by non-invasive brain stimulation techniques, such as transcranial direct current stimulation (tDCS; Coemans et al., 2021; Nissim et al., 2020; Norise & Hamilton, 2017; Roheger et al., 2024). TDCS can be used to modulate activity and neuroplasticity of target brain regions via weak electric currents either in an excitatory (anodal tDCS) or an inhibitory (cathodal tDCS) manner (Antal et al., 2017; Bikson et al., 2016; Nitsche & Paulus, 2000). Importantly, tDCS is well-tolerated and can be easily combined with behavioural treatment, making it a promising adjunct therapy for people with chronic post-stroke aphasia (Crosson et al., 2019; Fridriksson et al., 2019; Meinzer et al., 2016; Norise & Hamilton, 2017) or progressive neurodegenerative aphasia (Coemans et al., 2021; Cotelli et al., 2019; Roheger et al., 2024; Tsapkini et al., 2018).

Recent technological advances allow to safely deliver tDCS in home-based settings (Antonenko et al., 2022; Cappon et al., 2022; Charvet et al., 2015; Sandran et al., 2019). Several guidelines and recommendations have been published and suggest that home-based tDCS can be self- or caregiver-administered across age groups (Rocke et al., 2024) and diseases such as depression (Woodham et al., 2024), Parkinson’s disease (Dobbs et al., 2018), multiple sclerosis (Kasschau et al., 2016) and Alzheimer’s dementia (Satorres et al., 2023).

Together, these advances may enable the combination of tDCS and telerehabilitation to be administered in a home-based setting for pwPPA, thereby offering evidence-based and accessible treatment options. Despite numerous advantages (e.g., removing barriers to access in rural areas or to access to specialized but distant speech-language therapists, minimizing mobility requirements and traveling burdens and costs), this combined approach poses several (technological) challenges not only to rehabilitation researchers, but also to pwPPA and their caregivers. Therefore, it is important to involve these stakeholder groups in the development of such combined treatment approaches (Hersh et al., 2022).

Here, we used a two-stage iterative participatory approach, involving (1) semi-structured interviews and (2) usability tests to involve pwPPA and caregivers in the early phase of the development of a combined programme comprising home-based tDCS and a tele-adaptation of an established lexical naming training (Meinzer et al., 2016; Menke et al., 2009; Schomacher et al., 2006; Stahl et al., 2019) and communicative-pragmatic therapy (Breitenstein et al., 2017; Grewe et al., 2020).

We aimed to investigate the priorities and needs of pwPPA and their caregivers for treatment delivery and outcomes and to identify potential challenges and technological barriers related to the home-based delivery mode of SLT and brain stimulation. Using this approach, we sought to increase the overall acceptability and feasibility of the combined home-based tDCS and telerehabilitation approach by addressing needs, challenges and implementation ideas of pwPPA and their caregivers, and to facilitate future implementations in a clinical trial setting.

## Material & Methods

### Experimental procedure

We describe the two-stage, stakeholder-oriented development of a home-based tDCS and SLT telerehabilitation programme for people with PPA. In a first phase, semi-structured interviews were conducted to assess priorities and needs of pwPPA and their caregivers for telerehabilitation, SLT and tDCS, and to identify potential technological problems and barriers. Results of the interviews were used to develop a detailed step-by-step manual for combined tDCS and synchronous aphasia teletherapy in a home-based setting. In a second phase, this manual and its procedures were tested at the participants’ homes. Results of this usability test phase were used to refine a step-by-step manual. All methods and results are reported in accordance with the recommendations for reporting qualitative research (SRQR; O’Brien et al., 2014).

### Participants

Participants were recruited across Germany via the newsletter of the German Alzheimer Association and calls for participation that were distributed in memory clinics and outpatient practices for speech and language therapy. Eight pwPPA (age range: 58–82 years) and 16 caregivers (age range: 38–80 years) participated in two semi-structured interviews (see Tables 1 & 2 for participant demographics). This sample size is in line with recommendations for interviews according to which data saturation is reached between six to twelve interviews (Guest et al., 2006; Hennink & Kaiser, 2022). PwPPA were included when they had a neurologist-confirmed diagnosis of PPA (all variants), were able to communicate well enough to participate in an online interview format (assessed by trained speech language therapists (AR, NU)) and had no additional neurological or psychiatric diseases. There were no other exclusion criteria. Caregivers were included if they were family members or close friends of a person with PPA, even if the respective pwPPA were not eligible or willing to participate. In the usability tests, dyads consisting of caregiver and pwPPA were included. To make sure participants met all inclusion criteria, telephone screenings were conducted prior to study participation.

Four pwPPA and four caregivers who also participated in the interviews, agreed to participate in the usability test (see Table 1 for pwPPA’s demographics).

**Table 1.**
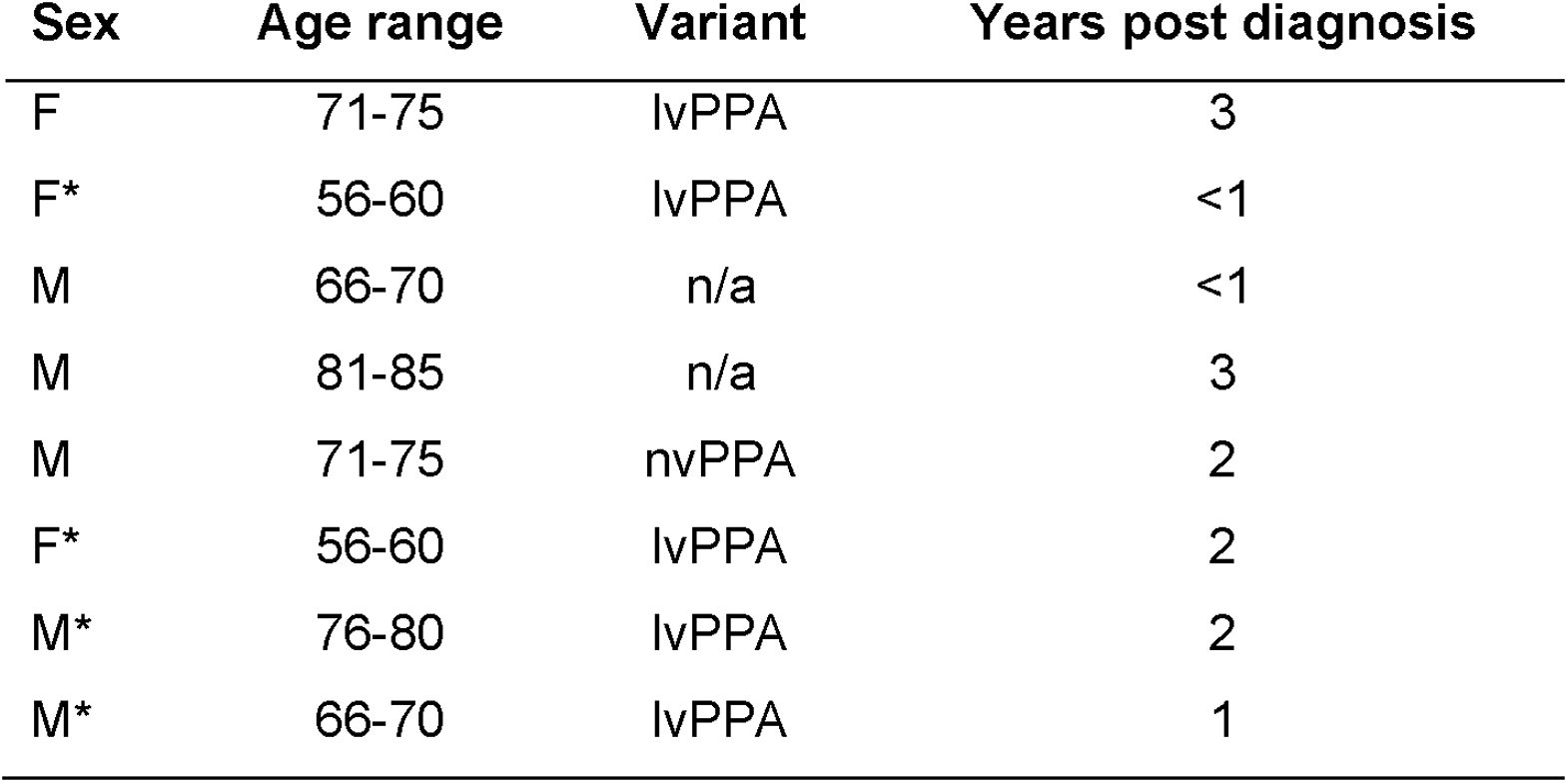
Demographic information of people with PPA who participated in the semi-structured interviews (N=8) and additionally in usability tests (N=4, marked with an asterisk). lvPPA = logopenic variant; svPPA = semantic variant; nvPPA = nonfluent agrammatic variant; n/a = PPA without further classification.

The study was approved by the local ethics committee (University Medicine Greifswald: BB 158/22, BB 196/23) and conducted in accordance with the Declaration of Helsinki. All participants provided written informed consent prior to the online interviews and/or before usability tests were conducted and were reimbursed with €10 per hour for their participation.

## Data acquisition

### Researcher characteristics and reflexivity

AR and RS conducted all interviews and usability tests. AR is a trained speech and language therapist. RS is a medical student. Neither AR nor RS knew the participants before the interviews but both knew half of the participants involved in the usability tests because of prior participation in the interviews. AR and RS developed the coding frame for the subsequent qualitative content analysis of the semi-structured interviews and analysed the semi-structured interviews. AR analysed the usability test data. A reflexive journal was kept by AR and RS during the data acquisition and analysis.

### Semi-structured interviews

Semi-structured interviews were conducted online via a secure teleconferencing platform (TrueConf; https://trueconf.com/), locally hosted by the University Medicine Greifswald. A twostage interview process was applied. In a first session, a short interview was conducted, focusing on general needs, challenges, and SLT-related priorities of pwPPA and carers (see Appendix Table 1 for an overview of the interview questions). This session was also used to check the availability and functionality of technical equipment (e.g., stable internet connection), to solve technical problems, and to refine the in-depth interview guide. The second interview (main interview) was conducted in a separate session and covered four broad topics: 1) speech and language therapy, 2) telerehabilitation (general), 3) non-invasive brain stimulation, and 4) the overall combined treatment approach (see Appendix Table 2 for an overview of the interview questions).

**Table 2.**
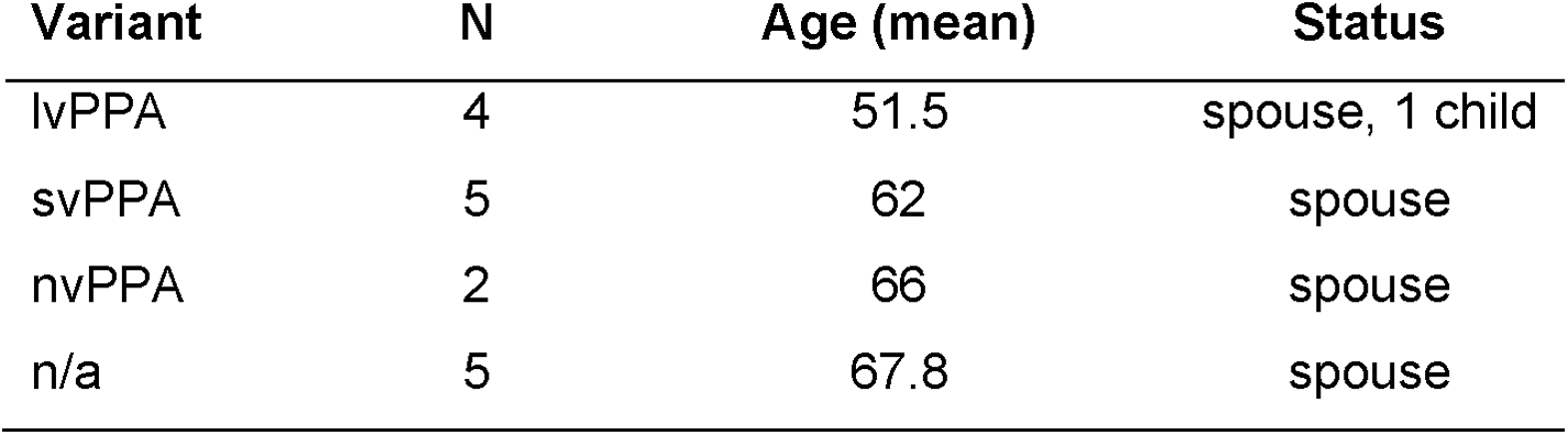
Demographic information of caregivers of people with PPA (N=16). lvPPA = logopenic variant; svPPA = semantic variant; nvPPA = nonfluent agrammatic variant; n/a = PPA without further classification.

Separate interviews were developed for pwPPA and caregivers. The broad categories were identical across the groups, but several specific questions differed (see Appendix Tables 1 and 2). The interviews comprised both open-ended questions as well as closed questions to enable qualitative and quantitative analyses.

All interview questions were embedded in a PowerPoint presentation and designed in an aphasia-friendly way, following the current guidelines for accessible information (Herbert et la., 2012). This involved the use of easier words, shorter sentences and illustrations with appropriate pictograms or figures to aid comprehension. Additionally, each question was presented on an individual slide. The PowerPoint presentation was shared via online screen-sharing. Interviews were recorded and subsequently transcribed by two trained researchers (AR and RS).

## Usability Tests

### Development of step-by-step manual

Selfor caregiver administration of tDCS and the ability to use a videoconferencing platform are important building blocks for the successful implementation of the planned combined home-based tDCS and telerehabilitation SLT approach. To investigate and ensure the feasibility of the home-based implementation of tDCS and the use of the videoconferencing platform, we developed a detailed step-by-step manual that was used to train patient-caregiver dyads (see Supplementary Material). BigBlueButton (https://bigbluebutton.org/) was chosen as videoplatform, because it is considered a secure teleconferencing platform, is hosted locally by the University of Greifswald and has all functions (e.g., an interactive whiteboard) which are necessary for the successful delivery of our planned speech-language therapy programme.

The usability of the developed step-by-step manual was documented and assessed by means of an established competency checklist that was adapted and extended for our purpose (Charvet et al., 2015; Cappon et al., 2023; see Supplementary Material).

Usability tests were conducted in the participant’s homes by two researchers (AR and RS) and followed a structured procedure to familiarize and train participants with the tDCS device and the telerehabilitation platform based on the step-by-step manual. The usability tests were split in three rounds per topic: In the first round, both researchers demonstrated the setup and handling of the tDCS device, by following the step-by-step manual. In the second round, the participant-caregiver dyads were asked to repeat the tDCS setup and handling using the stepby-step manual and the help of the researchers. Finally, in the third round, participants were asked to demonstrate the whole setup and tDCS handling independently, while this process was documented by one researcher using the competency checklist. The same procedure was repeated for the use of the telerehabilitation platform, followed by a short follow-up interview to inquire about additional challenges that were not captured by the competency checklists, and to document positive feedback about the usability of the manual, if applicable. Usability tests had a duration of three to four hours and were conducted during a single visit in the participants’ homes.

## Data analysis

### Semi-structured interviews

Interview data were analysed using a mixed-methods approach comprising qualitative content analysis (Kuckartz, 2018) and quantification (Vogl, 2017) in MAXQDA 2022 (VERBI Software, 2021). Closed questions were analysed quantitatively (percentages of responses), openended questions were analysed qualitatively. For the qualitative content analysis, coding frames were developed separately for both interviews in a deductive-inductive manner. The coding procedure involved two coding rounds. First, main categories were coded deductively with respect to the interview questions (short interviews) or broad categories (main interviews). Second, subcategories were identified within the main categories in an inductive manner. The coding frames were applied by two independent coders (AR, RS) and a subset of transcripts was coded by both coders to obtain a measure of intercoder reliability (Brennan & Prediger, 1981).

Next, category-based content analysis was performed within each group (caregivers, pwPPA; (Kuckartz, 2018; Rädiker & Kuckartz, 2019). To this end, participant-wise summaries were created for each category and subcategory. Finally, in order to identify commonalities and differences between the groups, we compared responses to overlapping questions between pwPPA and caregivers.

### Usability Tests

Data from the usability tests comprised competency checklists and brief follow-up interviews. The competency checklists were analysed quantitatively, using contingency tables. The brief follow-up interviews were transcribed by one trained person and analysed qualitatively using qualitative content analysis (Kuckartz, 2018). The coding frame for the brief follow-up interviews was developed deductively and followed the interview questions. The coding frame was applied by one coder (AR). Similar to the semi-structured interviews, category-based content analysis was performed within each group and subsequently compared between the groups (Kuckartz, 2018; Rädiker & Kuckartz, 2019).

## Results

### Results of the short interviews

In the short interviews, the themes were analysed along the questions concerning *Videoconferencing and PPA*, *Speech and Language Therapy* and *Telerehabilitation*. Three main themes emerged: 1) important general factors for videoconferencing, 2) SLT: important factors, expectations and desired outcomes, and 3) Telerehabilitation: important factors, expectations and desired outcomes. The intercoder reliability indicated a high level of agreement between coders (Kappa = 0.89; Brennan & Prediger, 1981).

#### Videoconferencing and PPA

Most caregivers indicated that pwPPA would need assistance to use a videoconferencing platform. Several potential challenges were identified. N=6 caregivers noted that pwPPA may lack technical experience, which can lead to excessive demands. One caregiver pointed out, that their relative with PPA had never worked with or used a computer on a regular basis. However, several caregivers argued that this problem might be a temporary problem, as digital literacy is increasing in the German society.

*Of course, we are currently in a generation where the people who are falling ill now are at an age where they have not yet grown up with technology, with the internet. That will of course change in later generations, when people have grown up with it. But in the current situation, this is still a challenge. (C_06)*

Also, some carers suggested that independent use of a videoconferencing platform may still be possible in early stages of PPA.

#### Speech and Language Therapy

Next, participants were asked about what is important to them in SLT and what expectations they have.

There was substantial overlap between pwPPA and caregivers with respect to priorities. Both groups named speech and language related topics as important factors. These included the reduction of word finding difficulties, improving their ability to read the newspaper or having conversations. One pwPPA (P_07) emphasized the general importance of SLT:

*“Speech therapy is important to me because it’s the only thing that does something to me.”* (P_07)

Four pwPPA emphasized the importance of regular SLT sessions. Apart from that, most pwPPA restricted their priorities to speech and language related factors, whereas caregivers added a broader range of priorities outside specific speech and language related topics. These included the previous experience of the therapist, frequency of therapy sessions, and provision of therapy early in the course of the disease.

Groups also differed in their outcome expectations. PwPPA expected an improvement or restoration of their ability to speak, whereas carers expected a slower rate of deterioration. However, all of them agreed that SLT is important for pwPPA

#### Telerehabilitation

Overall, both groups had an overall positive attitude, despite having little (N_pwPPA_ = 2/8) or no previous experience (N_pwPPA_ = 6/8) with telerehabilitation approaches. Most of the caregivers (N_carers_ = 14/16) were very enthusiastic about the possibility of telerehabilitation. An important argument for telerehabilitation was the increased availability and frequency of SLT sessions. Some caregivers (N_carers_ = 6/16) raised concerns that successful telerehabilitation depends on the quality of the internet connection and technical equipment. They also commented that inperson SLT, unlike telerehabilitation, also serves the purposes of personal social interaction with the therapists and other patients (in group therapy settings), facilitates communication by means of mimics and body language, and increases functional independence (i.e. traveling to the SLT independently). These facets may not be transferable via telerehabilitation.

### Results of the main interviews

The main interviews covered four broad themes: 1) speech and language therapy, 2) Telerehabilitation, 3) tDCS, and 4) the overall combined approach (see Figure 1 for an overview of the main four main themes and subthemes). There was a high level of agreement between coders as measured by intercoder reliability (Kappa = 0.85; Brennan & Prediger, 1981). Openended questions were analysed qualitatively and closed questions were analysed quantitatively (see Figures 2-4).

**Figure 1.**
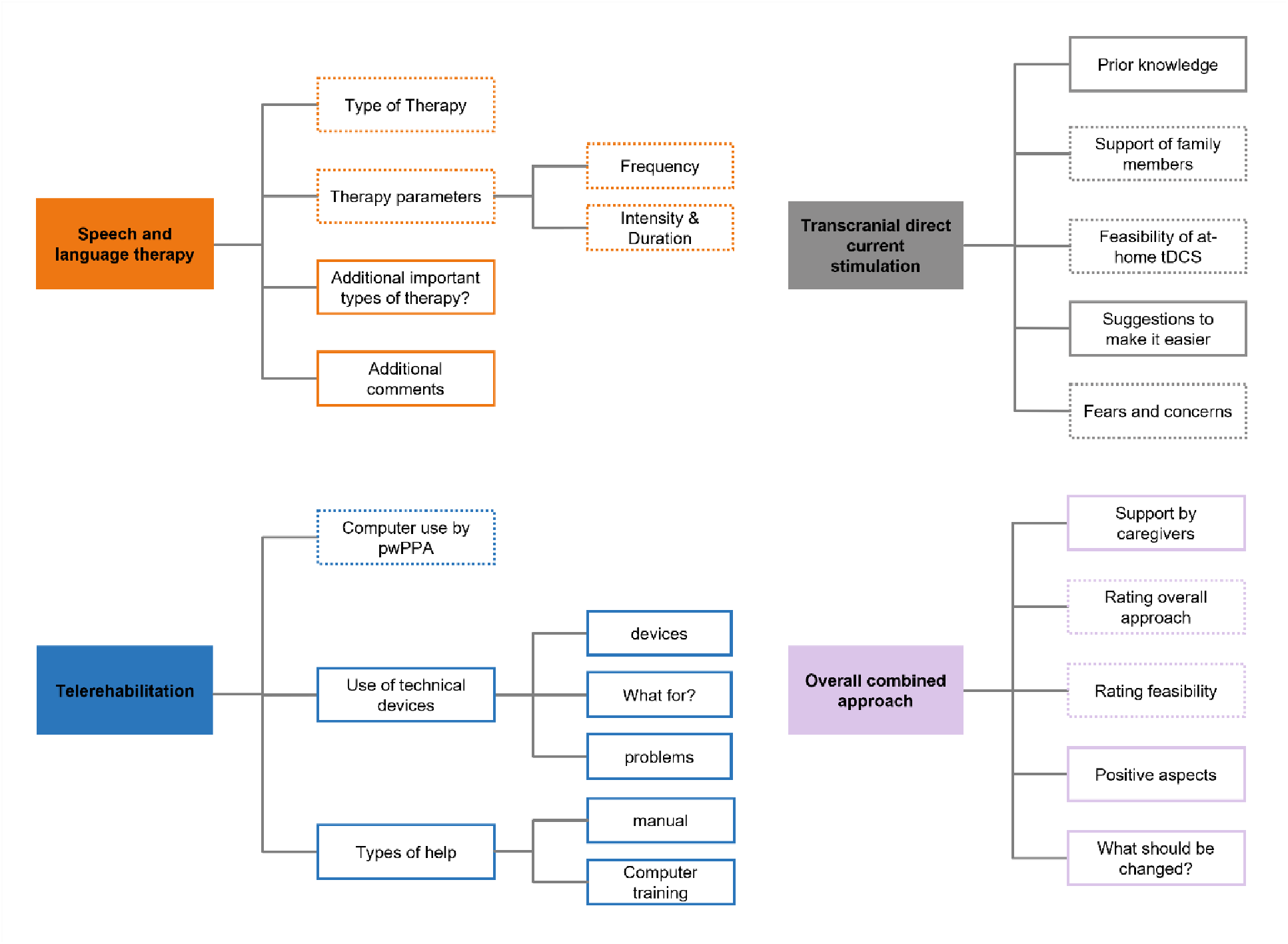
Coding tree for the main themes and subthemes of the main interviews. Main themes are illustrated in coloured rectangles, subthemes are displayed are colour coded respectively. Dotted outlines represent those subthemes that were analysed both quantitatively and qualitatively, solid outlines represent subthemes that were analysed qualitatively.

**Figure 2.**
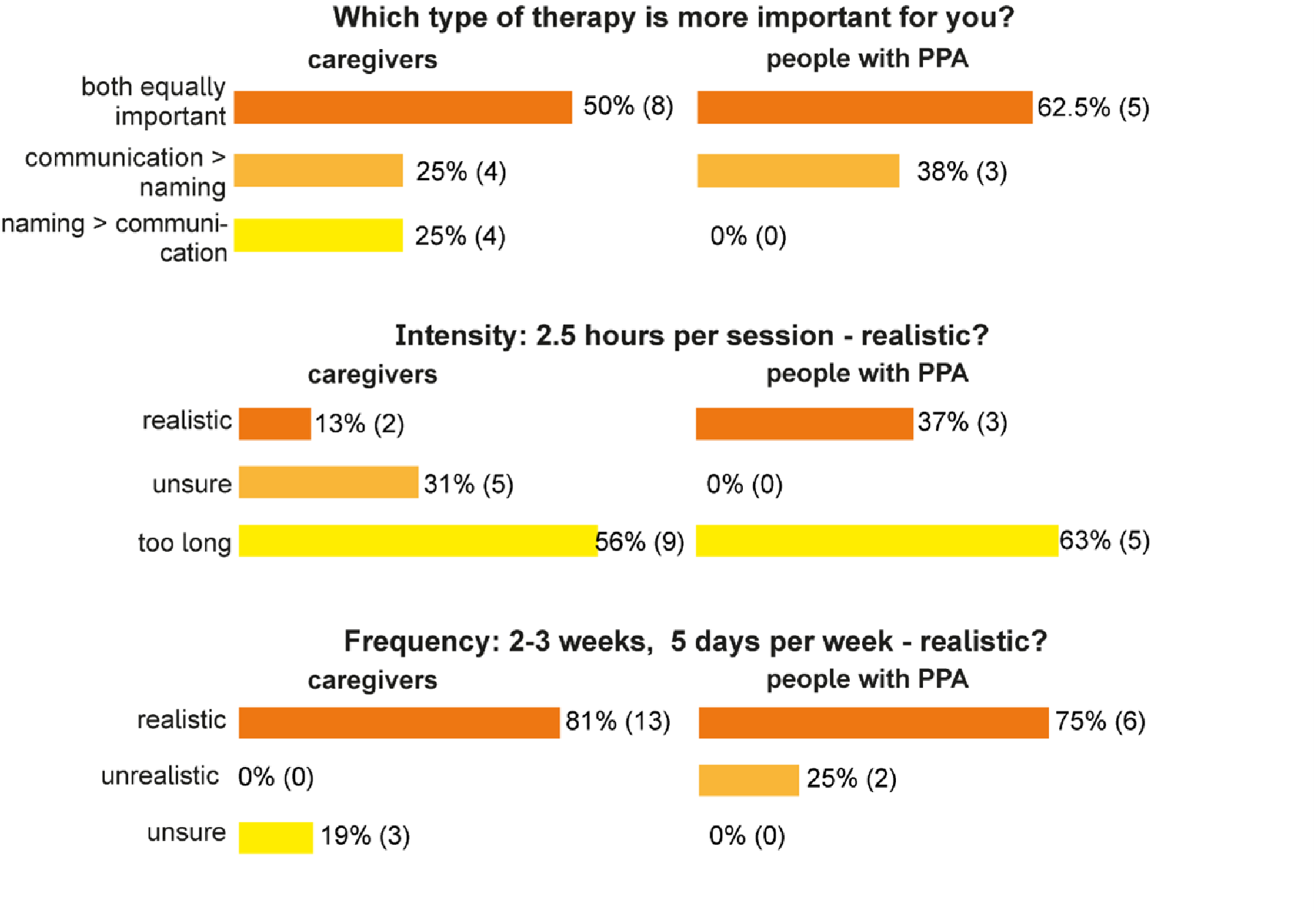
Quantitative results for the main theme Speech-Language Therapy. Upper panel: Caregivers (left) and pwPPA (right) prefer a combination of naming and communication therapy over one of both in isolation. Middel panel: Both groups indicate that an intensity of 2.5 hours per therapy session would be too long. Bottom panel: Both groups indicate that a daily treatment programme for two to three weeks would be realistic.

#### Speech and language therapy

Both groups had a clear preference for combining naming and communication training as opposed to a stronger focus on one of the two types of therapy (Figure 2, upper panel). N=3/8 pwPPA and N=4/16 carers responded that communication training was more important than naming:

*“Maybe even communication is even better I would say than naming therapy. Then getting in touch with people. I would say communication is more important I would say.” (P01)*

One caregiver (C_05) underscored the motivational aspect, which is higher in communicative settings than in computerized treatments:

*“The word-finding story [naming training], as in the example with the computer, has the weakness that it quickly becomes schematic. And that it quickly becomes boring and tiring at some point.” (C05)*

While 4/16 caregivers responded that naming was more important than communication training, none of the pwPPA shared this view. One argument that was mentioned by a caregiver in favour of the naming training was that it is relatively easy to create successful experiences and therefore to increase motivation:

*“I believe that it is easier to convey a personal sense of achievement with the naming exercises or naming therapy because it is simply less complex and there are many more opportunities for the patient to find confirmation and perhaps also to document or prove a learning effect.” (C06)*

Another caregiver raised the point that the most suitable type of therapy may change with disease progression:

*“So that depends very much on the stage of the disease. In other words, both are equally important at the beginning of the disease. Of course, the disease first becomes apparent when individual words are no longer present and cannot be assigned, found or interpreted. This means that word training is always important because it is the main difficulty at first. But as long as the patient is still at an early stage, this communication training can also be carried out well […] This means that the more advanced the disease, the more the focus will be on these pure word-finding therapies.” (C07)*

Another caregiver pointed out that incorporation of compensatory (e.g., non-verbal communication) strategies may become more important with disease progression and could be trained during communication training.

We also asked whether other types of therapy than naming or communication training were important for pwPPA and carers. Three caregivers mentioned music therapy and one caregiver suggested to incorporate naming and processing of numbers into speech-language therapy. Three caregivers mentioned that it may be relevant to train strategies for coping with stress in (unsuccessful) communicative situations. None of the pwPPA mentioned other types of therapy.

One pwPPA and two caregivers additionally pointed out that trained words and communication scenarios should be tailored to individual needs to increase motivation during therapy. Relatedly, one caregiver stressed that therapy materials are often designed for children or contain abstract sketches rather than realistic images. According to this caregiver “*patients have lost this ability to abstract, […] the images don’t have to be symbols, but actually depict the object as it looks in reality.*” (C07). Instead of suggesting another important type of therapy, one caregiver stressed that it was highly important to have regular therapy, as they had observed a deterioration of language functions after extended therapy breaks.

Next, we were interested in pwPPAs’ and caregivers’ opinions about the planned intensity and frequency of combined speech and language therapy (2.5 hours a day, 5 times a week for two weeks, for a total of 10 sessions). Most participants rated the planned intensity of 2.5 hours daily as too long (N_caregivers_=9, N_pwPPA_=5), but the frequency of daily treatment for a period of 2-3 weeks as acceptable (N_caregivers_=13, N_pwPPA_=6; Figure 2 middle and bottom panels).

Participants of both groups had alternative suggestions for the planned intensity and duration, ranging from 25 minutes (N_caregivers_=1), to half an hour a day (N_pwPPA_=1, N_caregivers_=1) to one hour a day (N_pwPPA_=1, N_caregivers_=3). Alternative suggestions included several breaks of flexible duration (N_pwPPA_=1, N_caregivers_=3), one long break of one hour (N_caregivers_=1), to split the therapy into a morning and an afternoon session (N_pwPPA_=1, N_caregivers_=5) or to decrease the daily intensity by increasing the overall therapy duration to 3-4 weeks (N_caregivers_=1). However, some participants favoured to have just a short break in between therapy blocks (N_pwPPA_=1, N_caregivers_=4).

Most caregivers and patients who considered the daily 2.5 hours as too long were concerned that it would be hard to concentrate for such a long period of time. One caregiver raised the issue that many pwPPA have a variety of (medical) appointments and that it may be difficult to fit intensive treatment regimes into personal schedules.

#### Telerehabilitation

With respect to the computer use by pwPPA caregivers gave mixed answers (Figure 3). Six caregivers stated that their relative with PPA was using a computer and had good technical abilities, whereas six other caregivers indicated the opposite. Those caregivers who were pessimistic about computer use by pwPPA reported poor digital skills prior to the onset of PPA, while those caregivers who were optimistic mentioned higher premorbid competency in smartphone and computer use. All eight participants with PPA used computers at home, although some preferred their tablet (N=2) or smartphone (N=4). Six pwPPA reported that they do not encounter problems or issues when using technical devices. Two pwPPA acknowledged that they occasionally encounter challenging situations, for example with complex passwords and that they ask their children or other relatives for support. Eight out of 16 caregivers stated that their relative with PPA was no longer able to use a laptop and that previous digital skills had deteriorated with progression of the PPA. PwPPA reported using technical devices for a wide range of activities such as communication (reading or writing emails, chat programmes, or other social media), entertainment (listening to music, watching videos, online shopping, booking flights, online banking), language or cognitive training (apps), or work. Caregivers reported broadly similar activities for their relatives with PPA.

**Figure 3.**
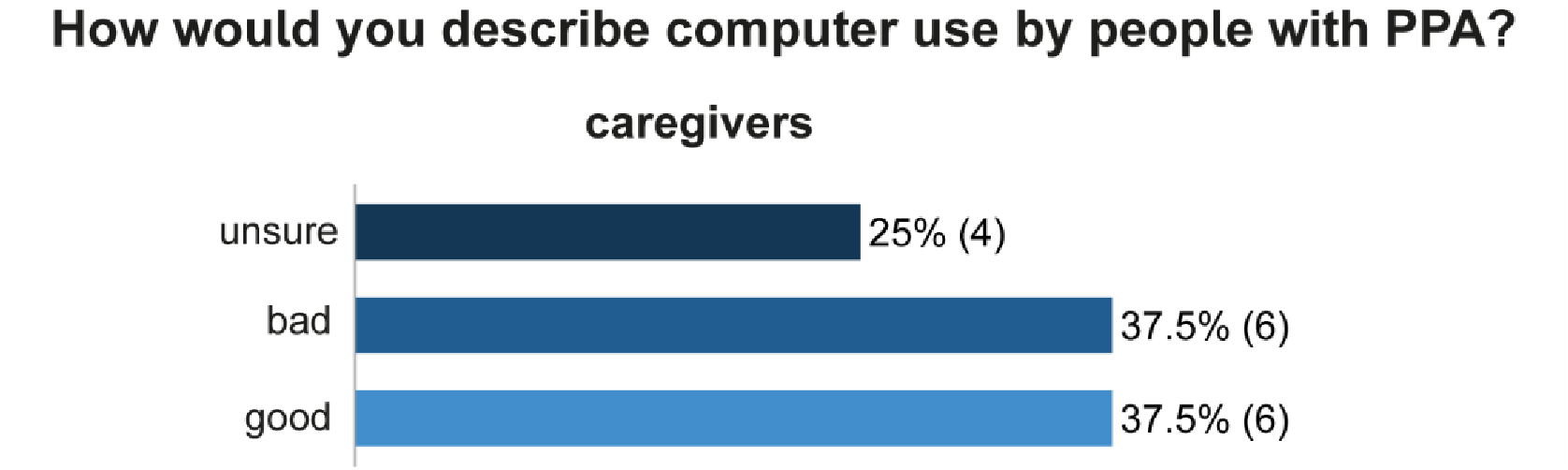
Caregivers’ perspective on computer use of pwPPA.

Next, we asked about common challenges that pwPPA experience when using a computer. Common problems encompassed both technical and participant-related domains. Technical issues included using a computer mouse or keyboard, difficulties with (complex) passwords, difficulties turning the computer on and off. Two caregivers commented that the use of a touchpad was more intuitive for their relative with PPA, and that this continued to work despite the inability to use a computer mouse. Participant-related issues included reduced problem-solving abilities, difficulty learning new skills and the refusal to ask for help when encountering difficulties:

*“So he’s not frustrated or anything. He tries again and again. And unfortunately it’s also the case that these people don’t necessarily call for help. […] In other words, sometimes I don’t even realise that he needs help.”* (C04)

Another caregiver emphasized that *“whenever something unforeseen happens, the pwPPA is usually faced with an unsolvable problem.”* (C07), demonstrating that pwPPA can successfully navigate familiar situations, but experience problems when they are outside of highly ritualised situations.

Finally, participants were asked about possible solutions to support pwPPA in using telerehabilitation. Most caregivers (N=12) indicated that computer training could be helpful. Important aspects for designing a computer training were “*to keep it as simple as possible*” (C16), to use “*very simple language, no foreign words”* (C07), and to choose “*the simplest possible device and the simplest possible operation of the operating system and software*.” (C07). Other suggestions were to use a tablet instead of a laptop, with *“large, [and] clearly illustrated touch surfaces”* (C07). Additionally, it was suggested to involve a caregiver in the computer training. In a similar manner, most caregivers indicated that a step-by-step manual with illustrations could be helpful. One caregiver pointed out that the use of pictograms in a manual would be too abstract and suggested to use real photographs with labels instead. Another caregiver underscored the importance of using large, easy-to-read fonts and simple sentences. However, caregivers differed in their opinion with respect to the level of detail that such an instruction or manual should contain. Some carers suggested to create a checklist that fits on a single page and contains only the most essential steps, while others suggested to illustrate each step.

#### Transcranial direct current stimulation

Approximately 50% of participants had previously heard of non-invasive brain stimulation (N_carers_ = 9/16, N_pwPPA_ = 3/8), yet most participants (N_carers_ = 10/16, N_pwPPA_ = 7/8) were optimistic about the use of tDCS and about the feasibility of independent self-administration or caregiver-assisted administration of tDCS at home (Figure 4). A number of suggestions were made to facilitate the handling and use of the tDCS device: having a single device to avoid misassembling different parts, simplifying the charging process, and labelling the devices and the button. Two caregivers said that a video tutorial would be helpful. Only 5/16 carers and 2/8 pwPPA had concerns or worries about tDCS and the specific concerns and worries were similar in both groups. Those included fear of pain or discomfort, lack of a desired effect, and fear of further brain damage or worsening of the condition. Most caregivers and pwPPA were open to the use of tDCS:

*“You have to dare to do something. I also take medication and I dare to swallow that little thing. I don’t know what’s in it either. I’m not worried about that either.”* (P07)

**Figure 4.**
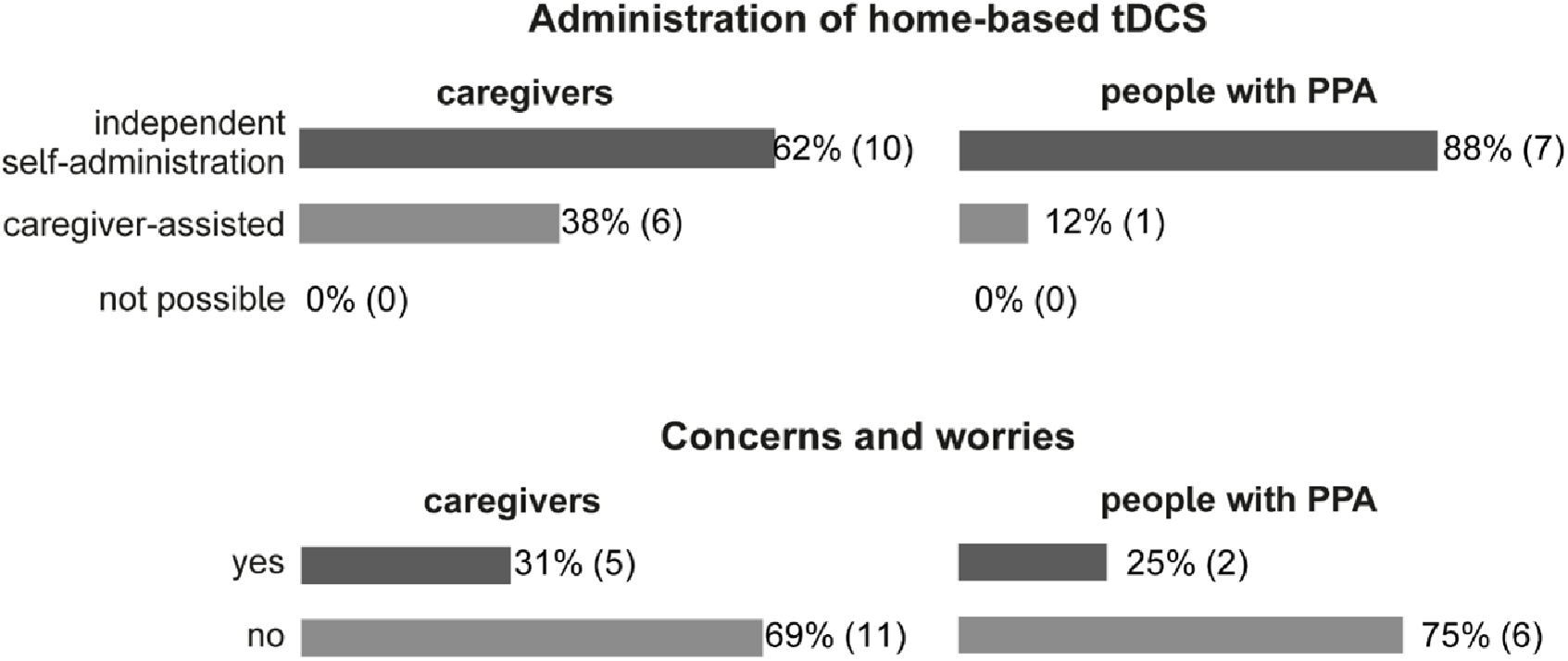
Results tDCS. The upper panel displays the feasibility rating of home-based tDCS administration, the lower panel shows the relative frequency of concerns and worries of caregivers (left) and people with PPA (right).

#### Combined telerehabilitation programme

Finally, we were interested in the participants’ opinion of the combined telerehabilitation programme. PwPPA and caregivers were asked to rate the overall concept and the feasibility of the concept on a scale from 1 (excellent) to 6 (bad). This scale is based on the German school grading system and was used to ease the rating for the participants. Both groups gave overall positive ratings, although caregivers gave a slightly better rating (mean rating caregivers: 1.5, mean rating pwPPW: 2.3; Figure 5 upper panel). Likewise, both groups rated the feasibility positively (mean rating caregivers: 1.9, mean rating pwPPA: 1.7; Figure 5 lower panel).

**Figure 5.**
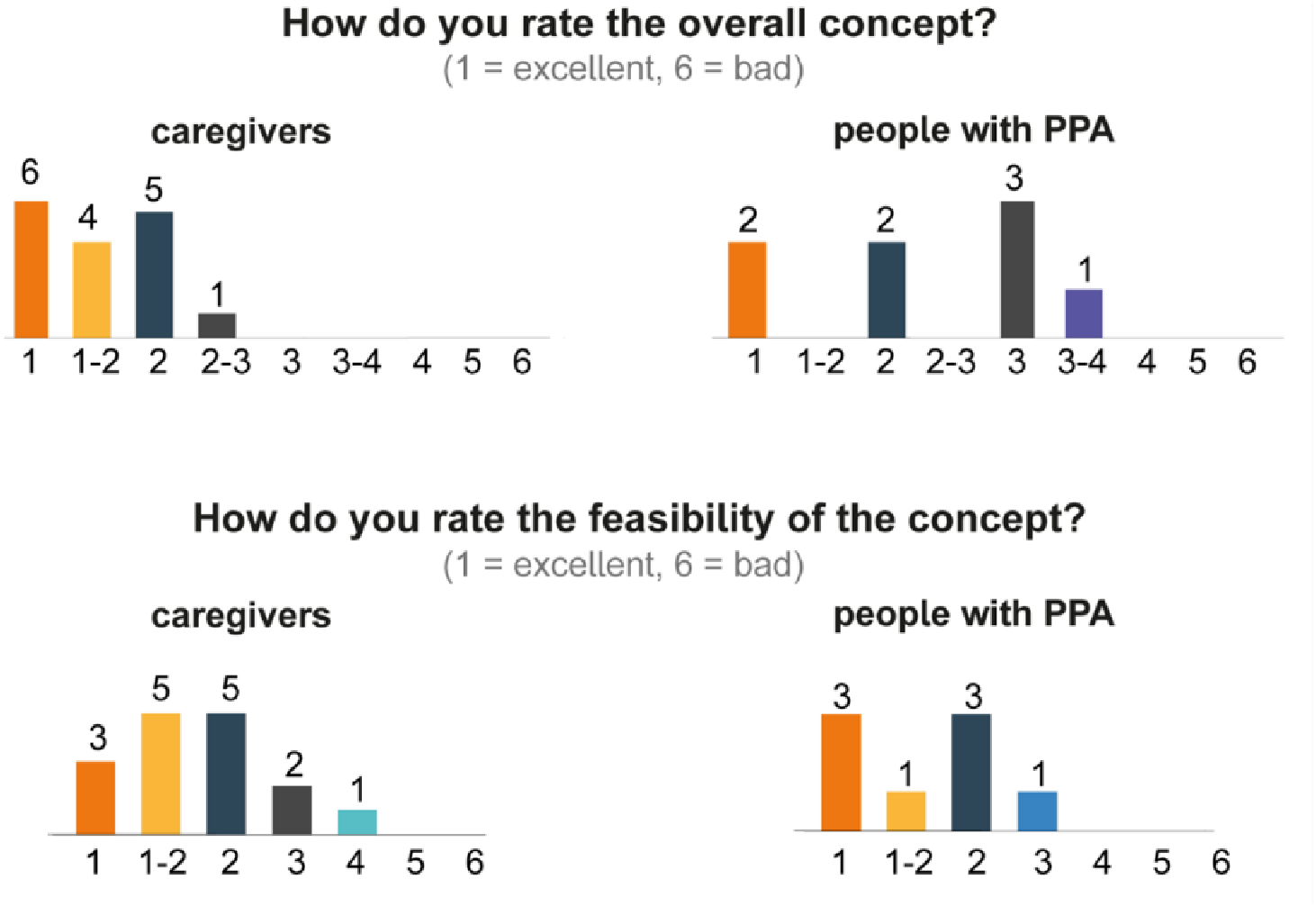
Quantitative results for the overall combined treatment approach. The upper panel displays the rating for the overall concept, the lower panel displays the rating for the feasibility of the overall concept.

Lastly, participants were asked to indicate what they would change about the planned treatment approach. The main criticism revolved around the planned intensity of 2.5 hours per day and was expressed by both groups:

*“Yes, I think the duration [intensity] might be a problem. So if we say we’ll do two one-hour sessions, I think that’s doable.”* (C08)

Another issue that was raised by two carers was the internet connection, which may be suboptimal in some parts of Germany. Another caregiver was concerned about the caregiver-support that the pwPPA would need during such a combined treatment and that it could pose an organizational problem for working caregivers. One pwPPA stated that they were concerned about the brain stimulation and that this would be their main reason not to participate in such a combined treatment approach. Similarly, one caregiver felt too uninformed and therefore concerned about tDCS. Another pwPPA reported that the combined approach was too complex and therefore hardly feasible.

We also asked, what participants liked about the treatment approach. As a positive factor several pwPPA and caregivers named the combination of brain stimulation and SLT:

*“So if I go to the speech therapist now and then I only have that one thing practically and then I have this thing on my head and so on. So that could be really good”.* (P02)

Two pwPPA also emphasized the novelty of the approach and potential benefits arising from the combination of SLT with tDCS. One pwPPA reported that what they liked most was the high intensity of speech-language therapy in such a short period of time.

One caregiver was keen about the development of a therapy programme itself:

*“I actually think it’s really good that there’s still something at all, because as I said, our initial information was that we couldn’t do anything and that’s a really bitter pill to swallow.”* (C04)

Finally, two caregivers mentioned the logistic benefits of telerehabilitation alongside increased accessibility as a positive aspect of the approach:

*“And that of course also saves resources and can help many people who perhaps don’t live in a city where there is a university hospital or something like that.”* (C03)

### Results of the usability tests

The results of the interviews were used to develop a step-by-step manual for the use of the telerehabilitation platform and the home-based tDCS setup. This manual and procedures were tested in the form of a usability test conducted in the homes of pwPPA and their caregivers. The results of the usability tests are summarized as actions performed either by (i) pwPPA or (ii) caregivers or (iii) pwPPA and caregiver together (Figure 6). Further information on usability was obtained by brief follow-up interviews, which were analysed qualitatively.

**Figure 6.**
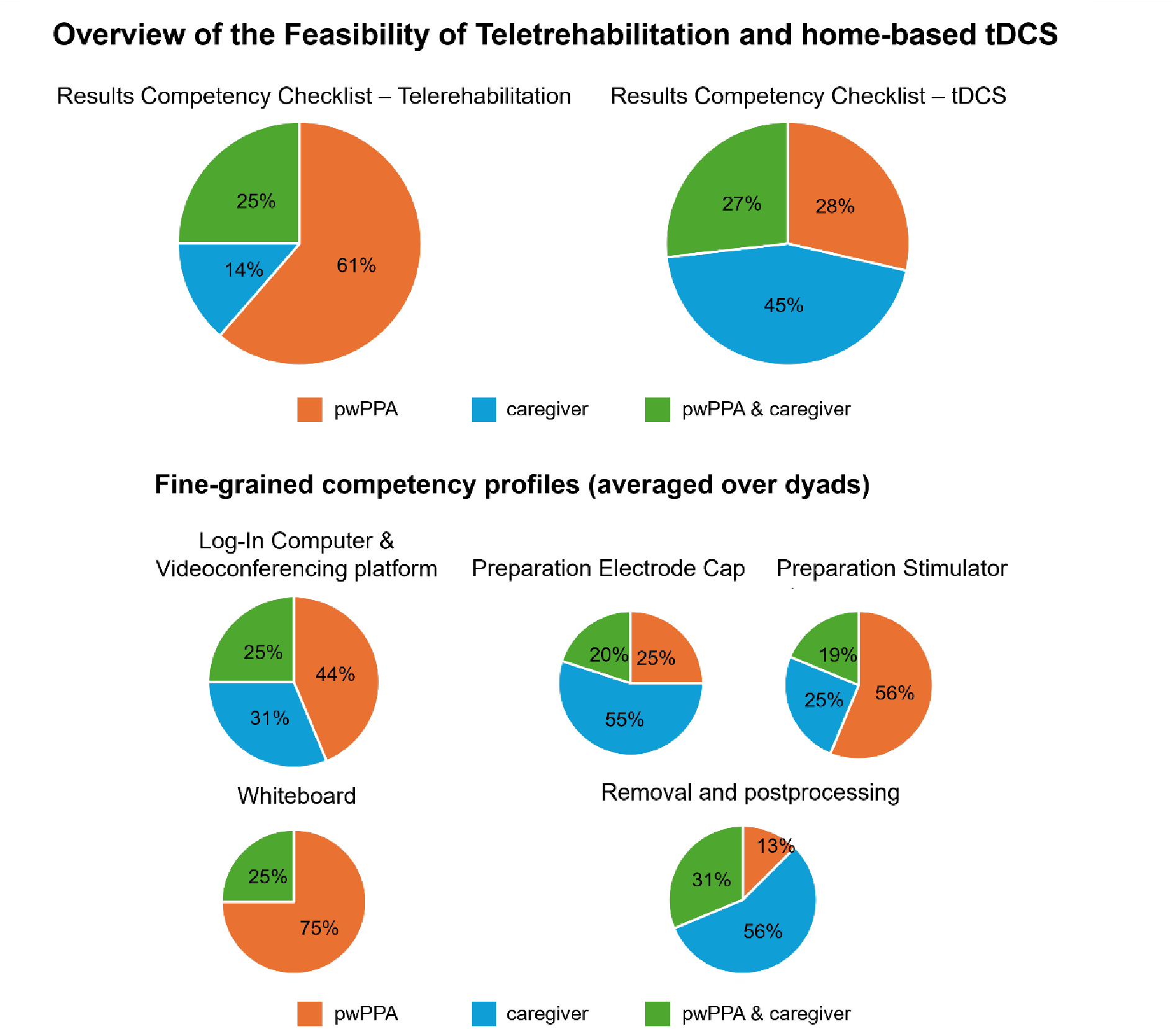
Feasibility of the individual steps of the step-by-step guide for telerehabilitation (left) and tDCS (right). Upper panel: Both figures show the mean percentage of actions that pwPPA were able to carry out independently (orange), pwPPA were able to carry out together with their carers (green) or that had to be fully carried out by caregivers (blue). Lower panel displays a fine-grained competency profile averaged over dyads, visualizing the subdomains of the competency checklists for telerehabilitation (left) and tDCS (right).

After training, all patient-caregiver dyads (PCD) were able to conduct all procedures that are defined in the competency checklist (see Figure 6 for an overview of the results of the competency checklists for telerehabilitation and tDCS and Appendix Figure 1 for an overview of the competency profiles of single patient-caregiver dyads).

Most of the actions (61 %) required to use the videoconferencing platform were performed by pwPPA alone. For 25 % of the required actions, pwPPA needed the assistance from their caregiver, and only 14 % of the actions were fully handled by caregivers. With regard to the use of the tDCS device, pwPPA were less independent. Caregivers executed nearly half of the actions (45 %) and assisted their relative with PPA in 27 % of actions. PwPPA were able to perform only 29 % of the required actions independently.

The follow-up interview covered three themes: positive aspects, difficulties and suggestions for the training procedure and step-by-step manual.

#### Positive aspects

The participants had a positive attitude towards the feasibility:

*“At first you think it’s a lot of steps, but because you do it repeatedly and more often, I don’t think it’s a problem for me now.”* (P_PCD_01)

*“I think the procedure is well described in the step-by-step manual, and you also showed us everything in great detail. The points to watch out for were again worked out in detail. Possible weak points or causes of error were also addressed. So, from my point of view, this is very feasible.”* (C_PCD_02)

The step-by-step manual was positively evaluated and well received by pwPPA and caregivers:

*“I think the explanations are very good, really foolproof. And I have no further questions about that. What I wanted to ask was explained very clearly.”* (P_PCD_04)

One pwPPA mentioned that although they found it more difficult to follow the instructions than her caregiver, it was feasible even for them.

#### Difficulties and suggestions for improvement

Two pwPPA stated that the most difficult aspect was the use of the whiteboard of the video conferencing platform. In contrast, the handling of technical devices (such as the tDCS device) was perceived as less difficult to use. One pwPPA reported that even the subjectively more difficult actions (e.g. handling of laptop and videoconferencing platform) were feasible when practiced.

With respect to the manual and general instructions, only one pwPPA experienced difficulties with reading as a consequence of their PPA. Therefore, this pwPPA focused on pictures in the manual instead of the text. Another pwPPA had difficulties reading the instructions and therefore suggested the use of larger fonts and larger images in the manual. Likewise, it was added that for some pwPPA it might be easier to follow the instructions when each step was printed on a single page.

Regarding the videoconferencing platform, one caregiver noted that screenshots on how to grant the software permission to use microphone and video were missing in the present version of the manual.

In the tDCS section, one caregiver found it difficult to insert saline solution into the electrodes without spilling it over the electrode cap. This caregiver suggested to highlight this in the manual, that one has to be very cautious. Several caregivers and pwPPA suggested to include additional notes for whether it is safe to wear glasses during stimulation and to exactly state when to put glasses on and off. It was further suggested by one pwPPA to provide replacement cables for the tDCS device, as the cables are very thin.

Finally, one caregiver recommended to conduct a mock session before the actual therapy sessions start to ensure that the overall technical setup and handling of devices work.

## Discussion

In the present study, we describe two stages of stakeholder-focused development of a homebased tDCS and telerehabilitation programme for people with PPA using semi-structured interviews and usability tests. The aim of the study was to evaluate the perspectives and potential (technological) barriers of pwPPA and their caregivers with regard to SLT, telerehabilitation and tDCS. The results of the first phase indicate that there is a high level of acceptance for intensive telerehabilitation SLT combined with tDCS. Based on these findings, a step-by-step manual was developed. The second phase demonstrated the usability of the developed manual and the feasibility of the technical aspects of the programme. In both phases, PwPPA and their caregivers provided valuable insights into all aspects of the treatment programme, which will inform further development, optimisation and implementation of the programme.

### Speech-language therapy

Both groups indicated a preference for a combination of naming and communicative therapy over either of the approaches as standalone therapy. This finding is in line with evidence suggesting that both types of therapy are effective for pwPPA (Jokel et al., 2014; Volkmer et al., 2020; Henry et al., 2019; Wauters et al., 2023). While the acceptance for the overall duration of 2-3 weeks was very high, both groups indicated that a daily intensity of 2.5 hours would be too long and exhaustive, although several participants of both groups indicated that it would be more acceptable with longer breaks. In addition, evidence from post-stroke aphasia demonstrates that self-reported fatigue during intensive therapy with a comparable treatment intensity is minimal (Pierce et al., 2024), suggesting that anticipated exhaustiveness may not necessarily correlate with perceived exhaustiveness. Moreover, it has been repeatedly shown that moderate-to-high treatment intensities provide significant treatment effects in post-stroke aphasia (Breitenstein et al., 2017; Rose et al., 2022; Brady et al., 2022), although evidence from PPA with respect to the optimal treatment dosage is currently missing.

### Telerehabilitation

The first stage (i.e., semi-structured interviews) identified several barriers with regard to telerehabilitation, including technical and participant-related issues.

For example, technical concerns identified by the caregivers included individual difficulties and preferences with the use of technical devices (computer mouse versus touchpad) and the lack of reliable internet connection in some regions of Germany. Both of these concerns can be addressed by providing appropriate equipment in a future clinical trial (i.e., laptops or tablets with the respective programmes and keyboard, mouse or touchpad, depending on the pwPPA’s preference), and by providing mobile Wi-Fi routers or internet sticks to ensure a fast and stable internet connection.

Several participant-related issues emerged, including the difficulty to learn new skills, reduced problem-solving abilities, the refusal to ask for help when difficulties are encountered, or impaired fine motor control. The difficulty to learn new skills, can be addressed through the (co-) development of appropriate telerehabilitation learning materials for pwPPA, which may help to increase motivation and accessibility. However, other identified participant-related issues, such as reduced problem-solving abilities or impaired fine motor control have been described in the literature before (Mooney et al., 2018) and can only be accounted for by caregiver involvement, which is a strategy that has been used in previous studies on telerehabilitation for pwPPA (Dial et al., 2019; Rogalski et al., 2016). This was reflected by the view of most caregivers, who indicated the necessity of a support person when using a computer. From the interviews, it became clear that not all pwPPA used computers or laptops, but some preferred smartphones or tablets, which may pose lower demands on (fine) motor skills or memory. Thus, it might be worthwhile taking these individual preferences into account: Telerehabilitation might be more acceptable if pwPPA are allowed to use their preferred technical device.

During the second stage (i.e., usability testing of the developed training and step-by-step manual), the use of the videoconferencing platform was described as challenging by pwPPA. This was likely due to the fact that pwPPA carried out 61 % of the actions independently. Caregivers supported 25 % of the actions and only 14 % were fully taken over, indicating that little support from caregivers was needed. Although described as challenging, these results indicate that it is feasible for pwPPA to navigate videoconferencing platforms and its tools with adequate and specific training. This finding is consistent with the results of a feasibility study on telerehabilitation for pwPPA, which identified the availability of a caregiver and prior familiarity with technical devices as facilitating factors (Rogalski et al., 2016) and with a recent single-case study in which a person with semantic variant PPA was trained successfully to use an app-based naming training on their smartphone (Joubert et al., 2024). Overall, the results of the usability phase demonstrate that pwPPA can effectively navigate videoconferencing platforms with appropriate training and minimal caregiver support, indicating the feasibility of teletherapeutic approaches.

### Transcranial direct current stimulation

The majority of pwPPA and their caregivers reported a positive attitude towards tDCS as a potential adjunct for SLT. However, tDCS is a relatively new method and consequently unknown to many people and it became evident that for those unfamiliar with tDCS, more information needs to be provided to enhance acceptability. In particular, layperson– and aphasia-friendly information about the mechanisms of action, the anticipated desired and adverse effects on brain function, and the excellent safety profile of the technique may be suited to reduce potential sources of fear and concern. This approach is accordance with recently published guidelines for the implementation of home-based tDCS, which underscore the importance of proper training of participants and assurance of their competency in the trained domains (Charvet et al., 2020) and the need for detailed written and video-based study-specific information materials (Antonenko et al., 2022). Moreover, these findings extend to information materials provided during the recruitment phase.

In the usability tests, the handling of tDCS was predominantly caregiver-assisted or fully managed by the caregiver. As a result, the use of the tDCS device was perceived as relatively easy compared to the use of the videoconferencing platform and whiteboard by pwPPA. However, it also shows that support by an additional person is required, when planning home-based tDCS interventions with pwPPA.

### Implications for future trials

All caregivers agreed that a step-by-step manual would be beneficial for pwPPA and also for the caregivers themselves, as they were required to assist their relative with PPA. This stepby-step manual would need to be designed in an aphasia-friendly way, i.e., with clear and unambiguous pictures that can be understood without the need for accompanying text, as pwPPA may experience difficulties with reading. Such a step-by-step manual has been developed based on the suggestions from the first stage and refined based on the suggestions from the second stage to ensure maximum utility (the final manual is available in the Supplementary Materials).

While pwPPA showed a high level of independence in the use of the videoconferencing platform and the whiteboard, caregiver-assistance was needed with respect to tDCS. Thus, in a future clinical study, home-based applications of tDCS for pwPPA should be planned in a caregiver-assisted and not fully self-administered manner.

## Limitations

While the online format of the interviews has the advantage of including participants who would not have been able to travel, it has some inherent limitations. First, there may be a bias towards including participants with a high levels of computer literacy or with supportive partners or family members. For example, pwPPA who lacked computer skills or who did not have supportive caregivers might not have been able to participate which may have biased the results towards high feasibility and acceptability of our approach. As PPA is a rare disease, which makes it very difficult to recruit participants, our sample consists of a small number of pwPPA, in the early stages of the disease. We were also not able to include pwPPA from all variants. Therefore, the reported views may not generalize to pwPPA in later stages of the disease or other subtypes of PPA. However, as participants did not necessarily have to participate as dyads, the views of caregivers of pwPPA in later stages and from more diverse subtypes could be included.

## Conclusion

In this mixed-methods study, we describe two phases of a participant-oriented development of home-based tDCS and speech-language telerehabilitation programme for people with PPA. Our findings contribute to a better understanding of the barriers, preferences and needs of pwPPA and their caregivers related to technically demanding therapy programmes. The results further demonstrate high acceptability for the planned programme and a high level of feasibility for the developed training procedures. Finally, the results highlight the importance of participatory research in the development and evaluation of new clinical interventions to ensure their acceptability and feasibility.

## Supporting information

Supplementary_Competency_Checklist

Supplementary_Manual

## Acknowledgements

The authors would like to thank the German Alzheimer Association, Prof. Dr. Dr. Matthias Schroeter, Frank Regenbrecht, Mirjam Gauch, Christina Molt and Lea Heidelmann for support with recruitment. We further thank all participants for taking part in the study.

## Author contributions

Conceptualization, A.U.R., M.M., and A.F.; Methodology, M.M., A.U.R., and M.R.; Investigation, A.U.R. and R.S.; Formal Analysis, A.U.R., R.S., M.M.; Resources, M.M. and A.F.; Data Curation, A.U.R.; Writing – Original Draft, A.U.R. and M.M; Writing – Review & Editing, A.U.R., T.G., N.U., C.B., F.B., A.F., M.M.; Visualization. A.U.R.; Supervision, M.M. and A.F.; Funding Acquisition. M.M., and A.F.

## Data Availability Statement

Video and audio data are not publicly available due to ethical reasons. Anonymized transcripts of interviews are available from the corresponding author on reasonable request.

## Funding information

This work was funded by the Federal Ministry of Education and Research (BMBF; Grant Number 01KG2210).

## Conflict of Interest Disclosure

The authors declare no conflicts of interest.

## Ethics Approval Statement

The study was approved by the local ethics committee (University Medicine Greifswald: BB 158/22, BB 196/23).

## Patient Consent Statement

All participants provided written informed consent prior to participation.

## Notes

### Competing Interest Statement

The authors have declared no competing interest.

### Author Declarations

Ethics committee of University Medicine of Greifswald gave ethical approval for this work (158/22, BB 196/23).

