## Supplementary_Material for "Participatory Development of a Speech-Language Telerehabilitation Intervention combined with Home-based Transcranial Direct Current Stimulation for Primary Progressive Aphasia: a Qualitative Study"

**Appendix Table 1.** Interview guide for the short interview.

| Theme | pwPPA | caregivers |
| --- | --- | --- |
| Icebreaker question | Did you have problems getting into this video conference? | Did you have problems getting into this video conference? |
| Videoconferencing & PPA |  | Can you imagine people with PPA successfully joining a video conference? |
|  |  | Do you have any ideas on how to make video conferencing more accessible for people with PPA? |
| Speech & Language Therapy | What is important to you in speech therapy? | What is important to you in speech therapy? |
|  | What outcome do you expect from speech therapy? | What outcome do you expect from speech therapy? |
| Telerehabilitation | Did you ever receive telerehabilitation, i.e. therapy via the internet? | Has your relative with PPA ever received telerehabilitation, i.e. therapy via the internet? |
|  | What is important to you in Telerehabilitation? |  |
|  |  | What do you think about telerehabilitation for people with PPA? |

**Appendix Table 2.** Interview Guide for the main Interviews.

| Speech and Language Therapy | pwPPA | caregivers |
| --- | --- | --- |
| Therapy Type | What’s more important for you? Do you think it is more important to find words again more easily (naming therapy) or to practise everyday situations (communication therapy)? Or do you find both equally important? | What’s more important for you? Do you think it is more important to find words again more easily (naming therapy) or to practise everyday situations (communication therapy)? Or do you find both equally important? |
| Intensity, duration & frequency | The speech therapy is planned to take place every working day for 2-3 weeks. Is that feasible for you? | The speech therapy is planned to take place every working day for 2-3 weeks. Do you think this duration is feasible for pwPPA? |
|  | Each session has a duration of approximately 2.5 hours. Is that feasible for you? | Each session has a duration of approximately 2.5 hours. Do you think this intensity is feasible for pwPPA? |
|  | Can you imagine taking part in 2.5 hours of speech therapy every day for 2-3 weeks? Is that feasible for you? | Can you imagine a pwPPA attending 2.5 hours of speech therapy every day for 2-3 weeks? Do you think this duration and intensity is realistic for pwPPA? |
| Telerehabilitation | **pwPPA** | **caregivers** |
| Computer competency | How do you feel about using a computer? (Scale from very confident to very insecure) | How do you rate the use of computers by people with PPA? |
|  | Do you have a stable Internet connection at home? |  |
|  | What do you use your computer for in everyday life? |  |
|  | What problems occur when using a computer? | What problems do pwPPA encounter when using a computer? |
|  | What difficulties do you see with speech therapy via the Internet? | What difficulties do you see with speech therapy via the Internet? |
| Tools & Aids | How can we ensure that you or other people with PPA can receive speech therapy over the internet? | Do you have any ideas on how to make it easier for pwPPA to use telerehabilitation services? |
|  | We can develop an illustrated step-by-step manual with pictures. We can offer you telephone support if technical difficulties arise. Do you have any additional ideas? |  |
|  |  | Can you imagine that computer training could help people with PPA to use telerehabilitation?  *Follow-up: How should such a computer training programme be structured? What do you think we should consider when planning a computer training programme?* |
|  |  | Can you imagine that an instruction manual with pictures could help people with PPA to use telerehabilitation?  *Follow-up: How should such a manual be structured? What do you think we should consider when creating a step-by-step manual?* |
|  | Do you need computer training to be able to use telerehabilitation? |  |
| Support for pwPPA |  | Can you imagine supporting your relative(s) with PPA in dealing with telerehabilitation? |
| tDCS | **pwPPA** | **caregivers** |
| Icebreaker Questions NIBS | Have you ever heard of brain stimulation or even received brain stimulation? | Have you ever heard of brain stimulation or even received brain stimulation? |
| Support for pwPPA | When carrying out stimulation at home, you will need support from a family member or other close carer.  Do you have someone who can help you? | When carrying out stimulation at home, the person with PPA needs support from a family member or other close carer.  Can you imagine that this is feasible? |
| Feasibility | We will provide you with training and instructions.  Can you imagine that brain stimulation can be feasible at home like this? | Can you imagine that after training and with step-by-step instructions, brain stimulation can be implemented at home? |
| Aids & tools | How can we make it easier for you? |  |
|  |  | Can you imagine people with PPA doing this on their own? Can you imagine supporting your relative with PPA to do this? |
| Worries & concerns | Do you have any worries or concerns about brain stimulation? | Do you have any worries or concerns about brain stimulation? |
| Overall treatment approach | **pwPPA** | **caregivers** |
| Overall concept | What grade (from 1=excellent to 6=poor) would you give the therapy concept? | What grade (from 1=excellent to 6=poor) would you give the therapy concept? |
| Feasibility | How do you rate the feasibility of the planned therapy concept? (from 1=excellent to 6=poor) | How do you rate the feasibility of the planned therapy concept? (from 1=excellent to 6=poor) |
| Positive aspects | What do you like about the therapy concept? | What do you like about the therapy concept? |
| Negative aspects | What would you change about the therapy concept? | What would you change about the therapy concept? |

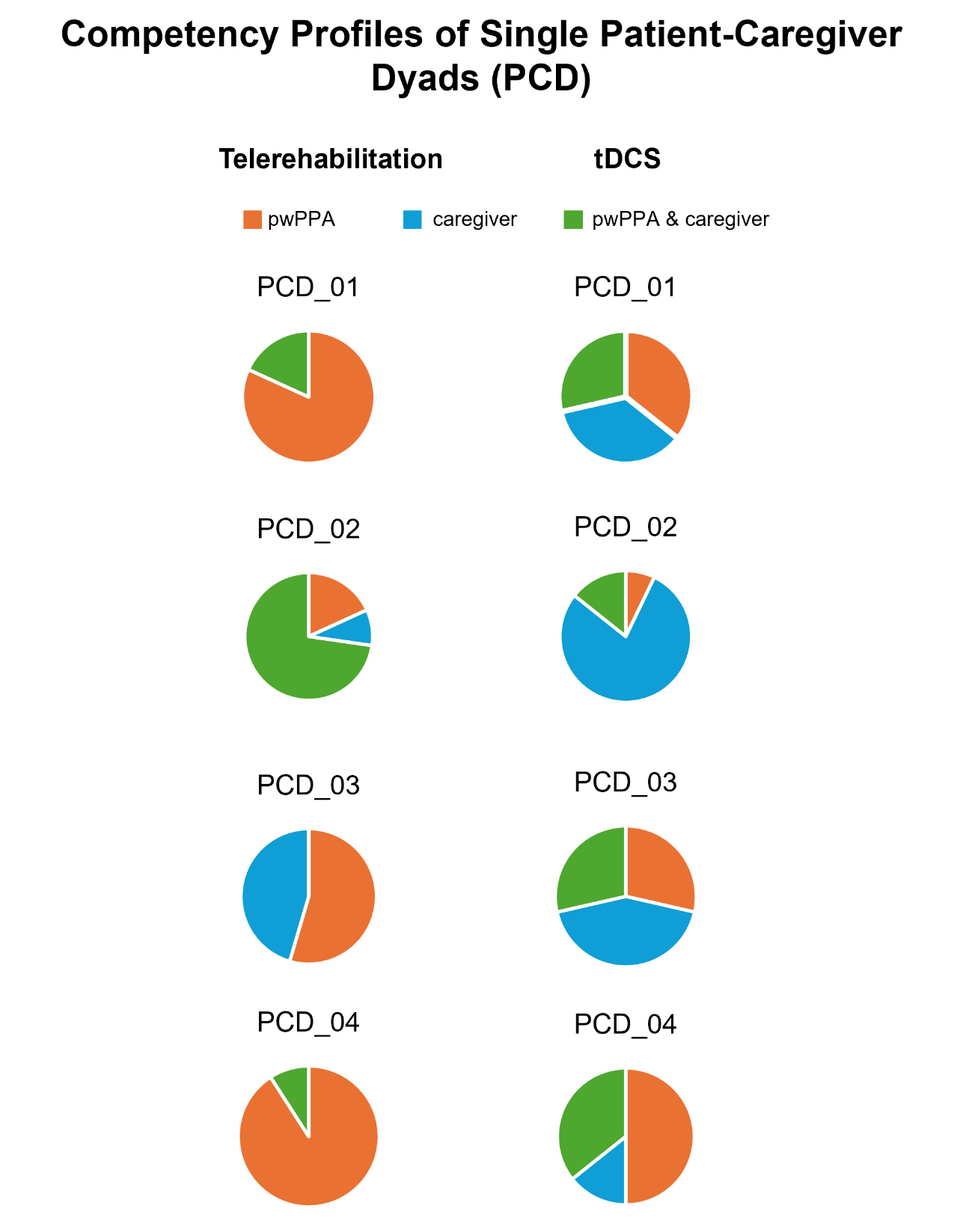

**Appendix Figure 1.** Overview of the competency profiles of single patient-caregiver dyads (PCD), based on the competency checklist results for telerehabilitation (left) and tDCS (right). Pie charts show the proportion of actions that pwPPA were able to carry out independently (orange), pwPPA were able to carry out together with their carers (green) or that had to be fully carried out by caraegivers (blue). These results illustrate the huge variability in competency profiles.
