## Supplementary_Competency_Checklist for "Participatory Development of a Speech-Language Telerehabilitation Intervention combined with Home-based Transcranial Direct Current Stimulation for Primary Progressive Aphasia: a Qualitative Study"

**Competency Checklist – HoBaT-PPA Phase IIIa – Training**

Participant ID:

Tester:

Date:

The following competences in handling the mobile tDCS device were demonstrated:

| **Competency** | **Carried out correctly?** | **Carried out by whom?**  **(C/P/C&P)** | **Comments** (e.g. questions, degree of assistance by caregiver or researcher) |
| --- | --- | --- | --- |
| **Knowledge of devices** | | | |
| Name all elements of the tDCS |  |  |  |
| **Preparations Cap** | | | |
| Check scalp for skin irritation |  |  |  |
| Check the surface of the cap and electrodes |  |  |  |
| Put on the cap correctly (end of cap should be directly above the eyebrows) |  |  |  |
| Fill each sponge electrode with 12 ml of saline solution using a plastic syringe |  |  |  |
| Attach electrode cable to cap (pay attention to colour-coding) |  |  |  |
| **Preparations Stimulator** | | | |
| Charging the memory module on the laptop |  |  |  |
| Connect electrode cables to stimulator (pay attention to colour-coding) |  |  |  |
| Connect stimulator to memory module |  |  |  |
| Start stimulation (theoretically) |  |  |  |
| **Follow-up work** | | | |
| Take off cap |  |  |  |
| Remove the electrode cable from the cap and stimulator |  |  |  |
| Wash the cap by hand with a suitable cleaning agent |  |  |  |
| Hang up the cap to dry |  |  |  |

The following competences in dealing with BigBlueButton were demonstrated:

| **Competency** | **Carried out correctly?** | | **Carried out by whom?**  **(C/P/C&P)** | **Comments** (e.g. questions, degree of assistance by caregiver or researcher) |
| --- | --- | --- | --- | --- |
| **Log-In** | | | | |
| Call up e-mail inbox |  |  | |  |
| Join the video conference via the invitation link and password |  |  | |  |
| turn on camera |  |  | |  |
| Turn on microphone |  |  | |  |
| **BigBlueButton Functions** | | | | |
| Find chat |  |  | |  |
| Write in the chat |  |  | |  |
| Turn whiteboard on and off |  |  | |  |
| **Whiteboard** | | | | |
| Write (text field) |  |  | |  |
| Draw (line) |  |  | |  |
| Draw (rectangle) |  |  | |  |
| Move object or text field |  |  | |  |
| Change the size of a text field |  |  | |  |
| Usethe eraser function |  |  | |  |

The dyad has successfully acquired all the skills required to use the mobile tDCS device.

o Yes

o No, a new competency check is required

The dyad has successfully acquired all the skills required to use BigBlueButton.

o Yes

o No, a new competency check is required

Name: _______________________________________________

Date: _____________________________________________

Additional comments: ____________________________________________________________________________________________________________________________________________________
