## Supplementary material for "Participatory Development of a Speech-Language Telerehabilitation Intervention combined with Home-based Transcranial Direct Current Stimulation for Primary Progressive Aphasia: a Qualitative Study": Tables

**Table 1.** Demographic information of people with PPA who participated in the semi-structured interviews (N=8) and additionally in usability tests (N=4, marked with an asterisk). lvPPA = logopenic variant; svPPA = semantic variant; nvPPA = nonfluent agrammatic variant; n/a = PPA without further classification.

| Sex | Age range | Variant | Years post diagnosis |
| --- | --- | --- | --- |
| F | 71-75 | lvPPA | 3 |
| F* | 56-60 | lvPPA | <1 |
| M | 66-70 | n/a | <1 |
| M | 81-85 | n/a | 3 |
| M | 71-75 | nvPPA | 2 |
| F* | 56-60 | lvPPA | 2 |
| M* | 76-80 | lvPPA | 2 |
| M* | 66-70 | lvPPA | 1 |

**Table 2.** Demographic information of caregivers of people with PPA (N=16). lvPPA = logopenic variant; svPPA = semantic variant; nvPPA = nonfluent agrammatic variant; n/a = PPA without further classification.

| **Variant** | **N** | **Age (mean)** | **Status** |
| --- | --- | --- | --- |
| lvPPA | 4 | 51.5 | spouse, 1 child |
| svPPA | 5 | 62 | spouse |
| nvPPA | 2 | 66 | spouse |
| n/a | 5 | 67.8 | spouse |
