## Supplementary_Manual for "Participatory Development of a Speech-Language Telerehabilitation Intervention combined with Home-based Transcranial Direct Current Stimulation for Primary Progressive Aphasia: a Qualitative Study"

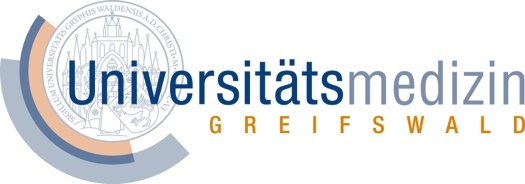

**Step-by-step manual**

**Transcranial direct current stimulation and teletherapy**

as part of the study

**"Home-based speech therapy and transcranial direct current stimulation in primary progressive aphasia"**

**Principal investigators:** Prof. Dr. Marcus Meinzer, Prof. Dr. Agnes Flöel

**Contact:** Anna Rysop

### 1. Introduction

Dear study participant,

This instruction manual is intended to prepare, perform and follow up on transcranial direct current stimulation ('tDCS') as part of the study 'Home-based speech therapy and transcranial direct current stimulation in primary progressive aphasia'.

In the following, all actions are presented and illustrated step by step. First, a brief device overview (Chapter 2) is given. Please read everything carefully and familiarize yourself with the devices. This chapter is followed by a description of the individual actions, divided into preparation for stimulation and therapy (Chapter 3), implementation of stimulation and therapy (Chapter 4), and follow-up of stimulation and therapy (Chapter 5). For a better overview, these steps are color-coded on the respective side margins. In between, you will find warnings highlighted in red and general information highlighted in green. Please read all of them carefully. Chapter 6 presents basic functions of the video platform BigBlueButton (BBB for short). BigBlueButton is used as a video platform for teletherapy as part of our study. Finally, in Chapter 7 you will find a collection of "Frequently Asked Questions" and related solutions.

Note that this step-by-step guide was prepared specifically for the study "Home-based speech therapy and transcranial direct current stimulation in primary progressive aphasia". If you have any further questions about the device or transcranial brain stimulation, please contact us or read the manufacturer's instructions for use.

### 2. Device overview

#### 2.1. Laptop und Charging Cable

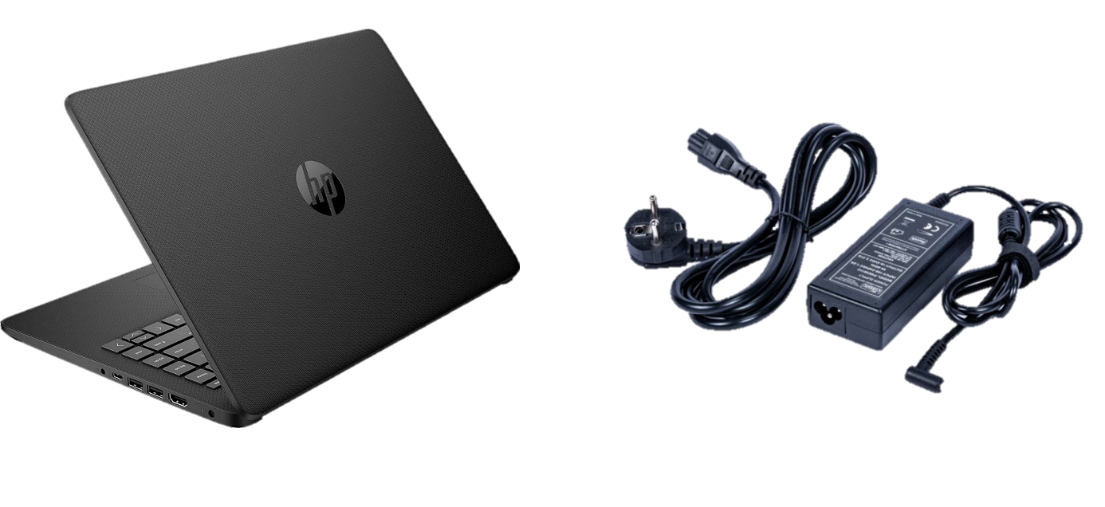

**Figure 1:** Laptop und Charging Cable.

The laptop is required for the following steps:

- Charging the stimulator
- Participation in teletherapy
- Transmitting the stimulation data

#### 2.2. Stimulator and components

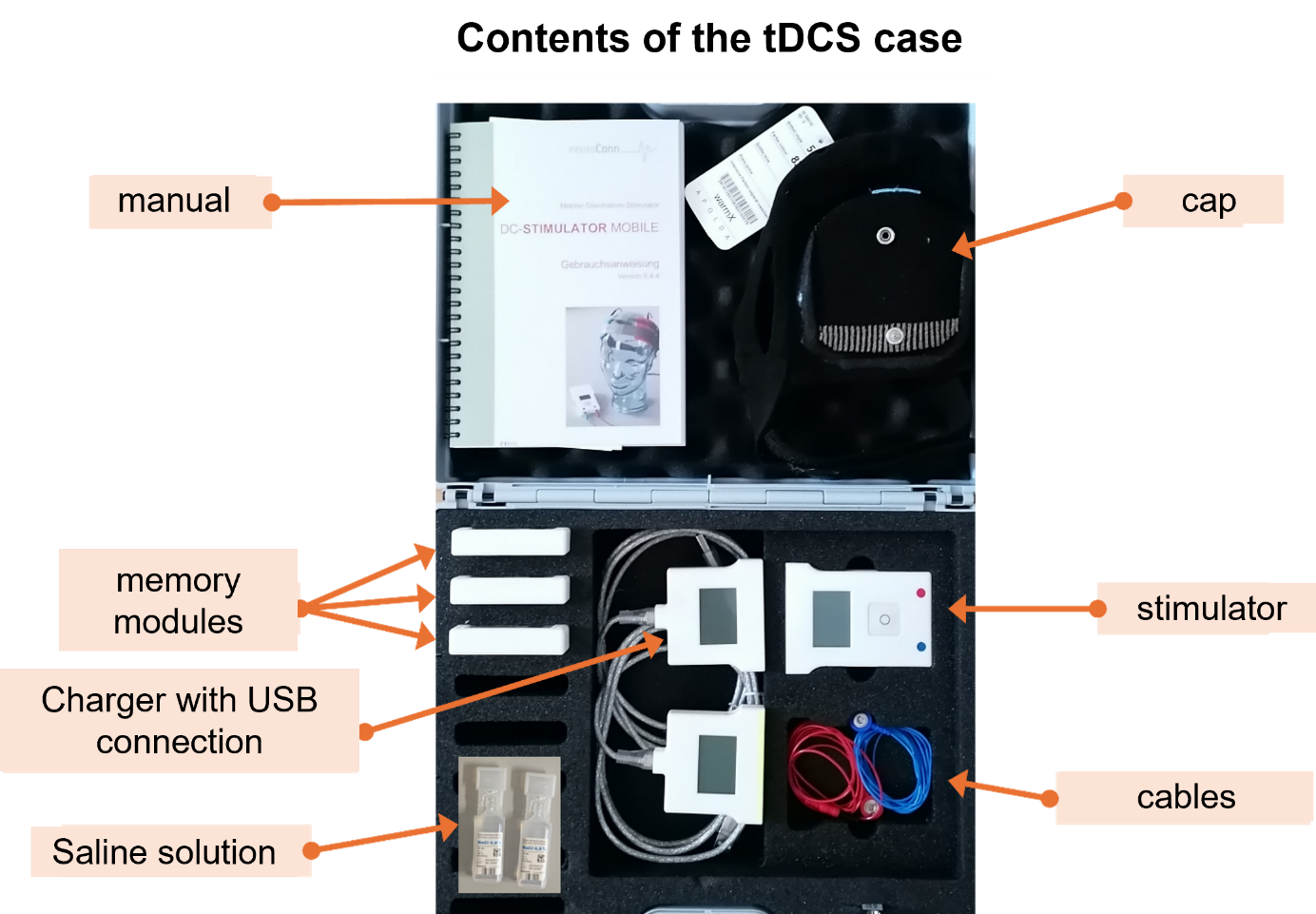

**Figure 2:** Contents of the tDCS case.

The stimulation kit contains all the necessary components for brain stimulation (see Figure 2 for an overview of the components included). Here's what's included:

- Manufacturer's instructions for use
- 1 cap including electrodes
- 3 memory modules
- 1 charger with USB connection cable
- 1 Stimulator
- 1 blue and 1 red electrode cable each
- Saline solution (NaCl)
- Syringe without cannula (not shown in Figure 2)

### 3. Preparations for stimulation and therapy

#### 3.1. Charging the devices

For successful participation in the therapy, you need the **laptop** and the **brain stimulator**. Please charge both devices in time.

The laptop can also be charged during therapy. The brain stimulator must be fully charged before therapy.

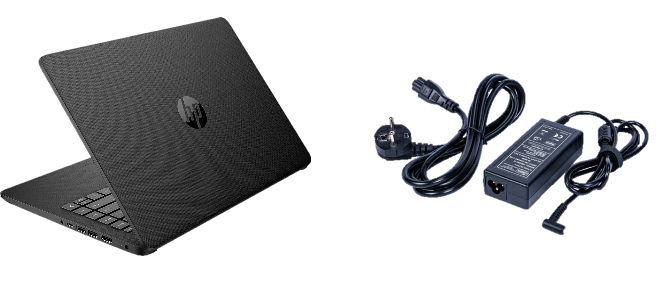

##### 3.1.1 charging the laptop

Required equipment:

- Laptop
- Charging cable

**Charging cable**

**Laptop**

To charge the laptop, connect the laptop to a power outlet. Plug the cable into the appropriate opening on the right side of the laptop. Now plug the power plug into a socket. When everything is plugged in correctly, a small orange lamp will light up on the right side.

##### 3.1.2 Charging tDCS Device

Required equipment:

- Memory module
- Charger (yellow marking) with USB connection cable
- Laptop

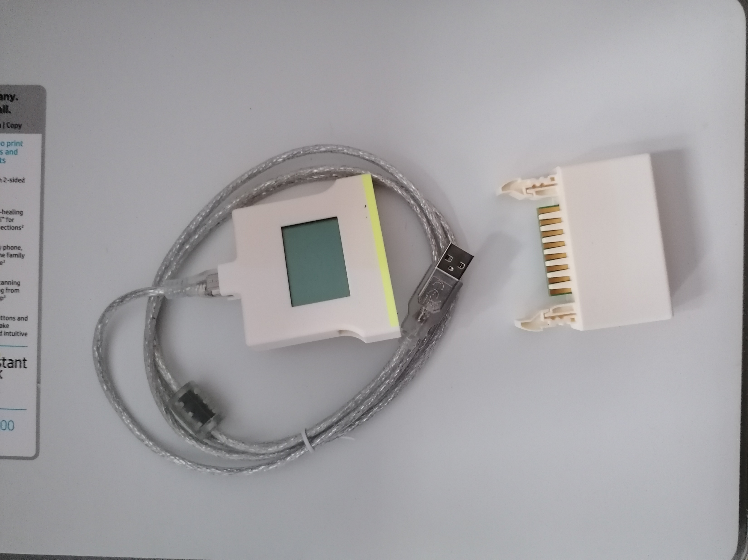

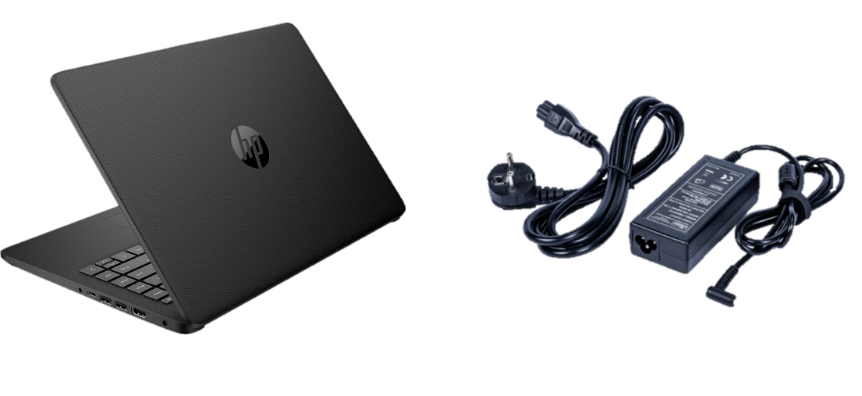

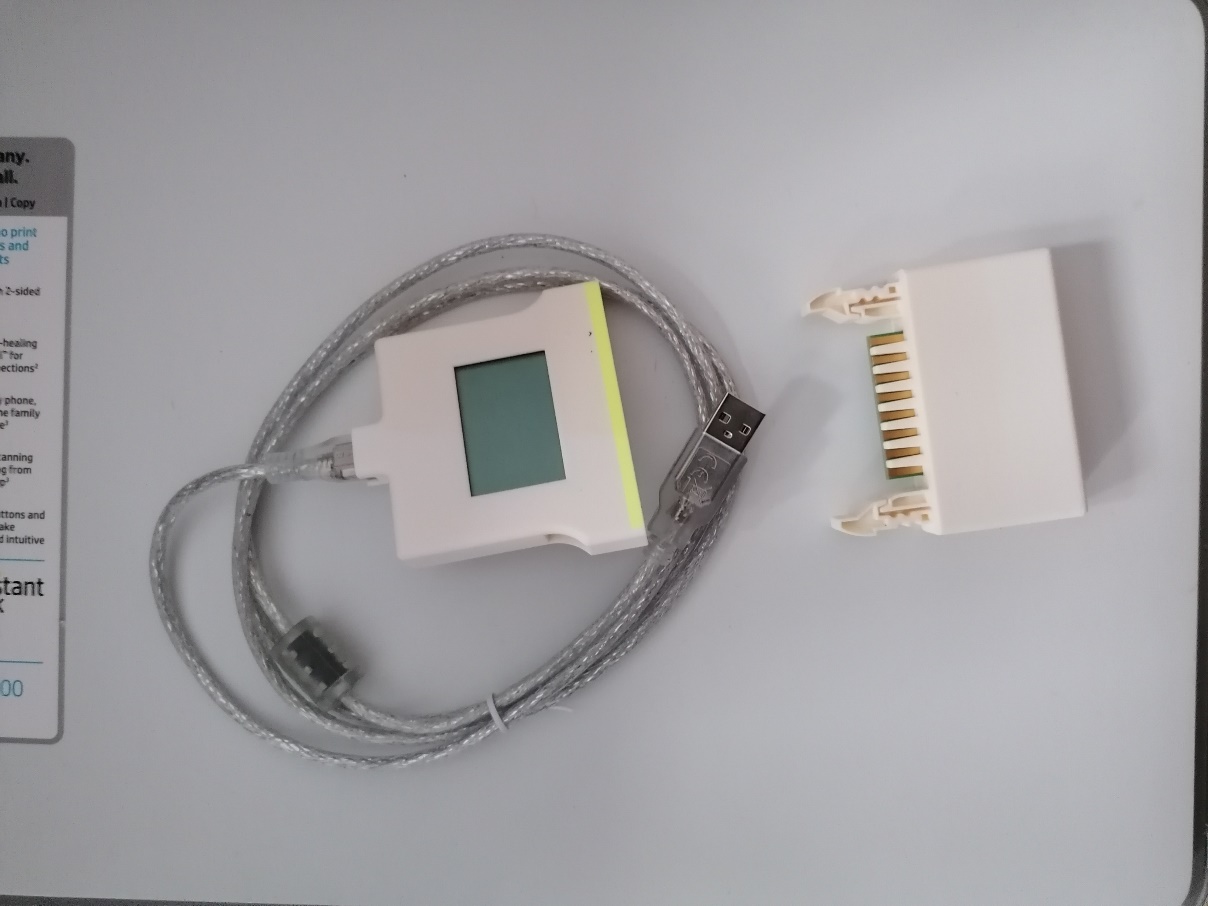

**Memory module**

**Laptop**

**Charger**

**Here's how to do it:**

| 1) Connect the **memory module** to the **charger**. To do this, take the memory module in one hand and the charger in the other. Make sure that the front of both devices is facing you. Plug both devices into each other. You should hear a **click**. | 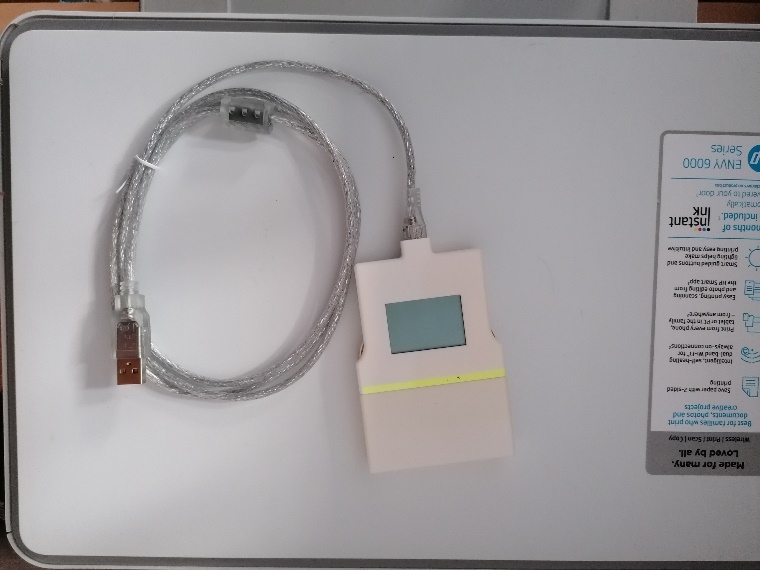 |
| --- | --- |
| 2) Connect the **charger** to the **laptop**. To do this, plug the **USB connection cable** of the charger into the USB input of the laptop. The laptop must be switched on for this. If you have connected everything properly, the screen of the storage device will light up. | 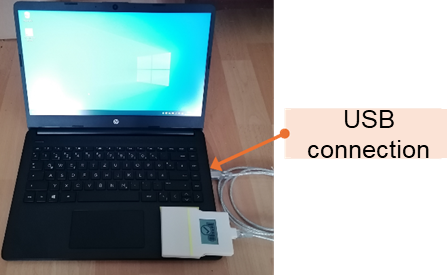 |
| 3) Keep charging the **storage device** until you see a fully charged battery and a check mark on the storage device screen.   It takes about 20 minutes **to fully charge**. | 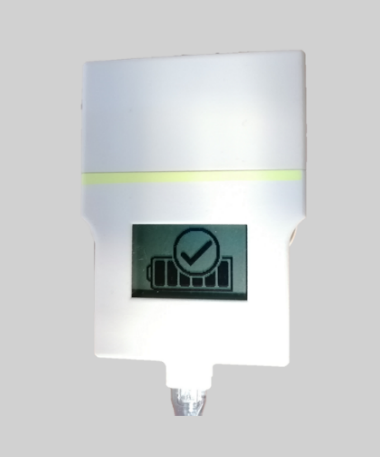 |
| 4) When the **storage device** is fully charged, unplug the **USB connection cable** from the **laptop**. | 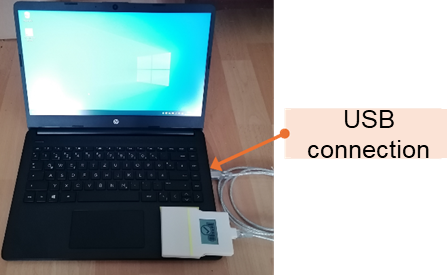 |
| 5) Disconnect the **storage device** and **charger**. To do this, squeeze the two side locking springs together and at the same time carefully pull out the memory module. | 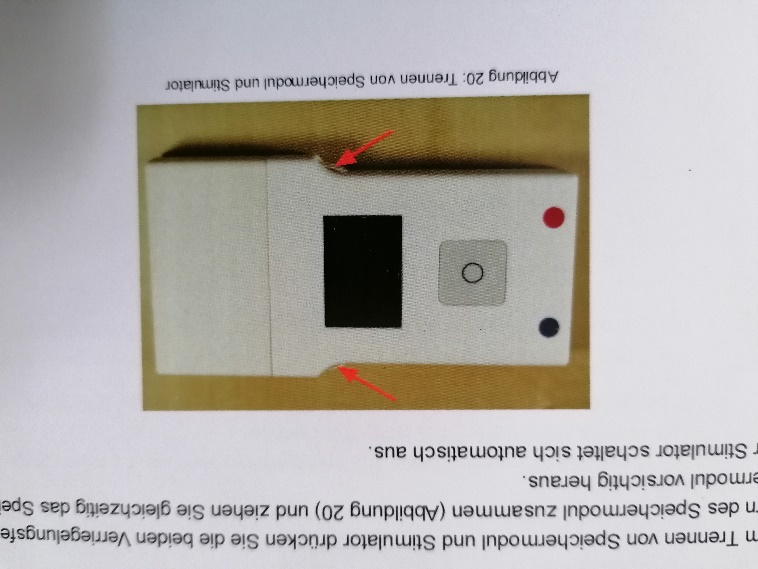 |
| 6) Now stow **all the devices** back in the **stimulation case** | 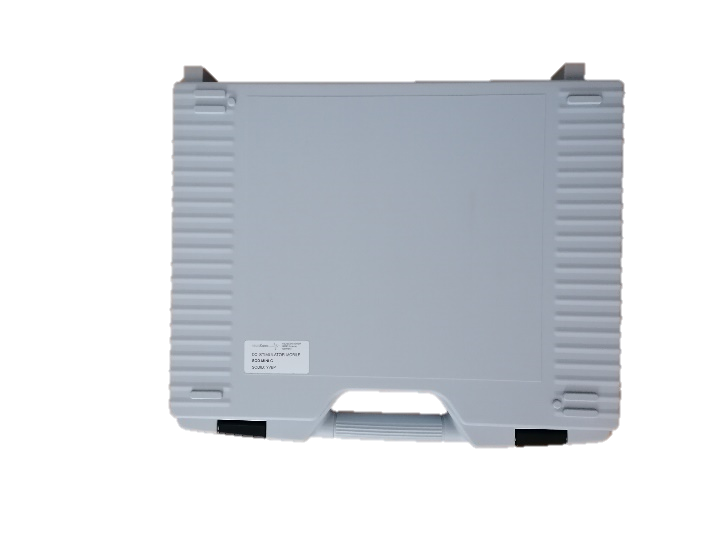 |

**Important:** The **memory module** must be charged daily. It is best to charge the memory module directly after the last therapy session of the day.

#### 3.2 Preparation for brain stimulation

Please prepare all necessary items.

Required items:

- Stimulator
- Memory module
- cap (dry)
- Red and blue electrode cable
- Saline solution (NaCl)
- Syringe (without cannula)

**Important:** The **hood** must be **dry**. If the hood is still damp in some places after a possible cleaning, you cannot start stimulation. Wait until the hood is **completely dry**.

**Please check that you have all the equipment you need in front of you before proceeding with the next steps.**

| 1) Check the condition of the **cap**.   The cap must **be dry**.  The cap must **be intact**. There should be no holes or cracks. | |
| --- | --- |
| 2) Check your **hair**. The hair must **be dry**. There must be no hairspray, gel or residues of other care products, such as foam or oil, on the hair. | |
| 3) Check the **scalp** in the areas where the electrodes rest. There should be no skin irritation or sores. | |
| 4) Put on the **cap**. To do this, pull the openings over the ears. The front part of the cap should end just above the eyebrow. The electrode marked in red should be on the left side of your body, the electrode marked in blue on your right side. | 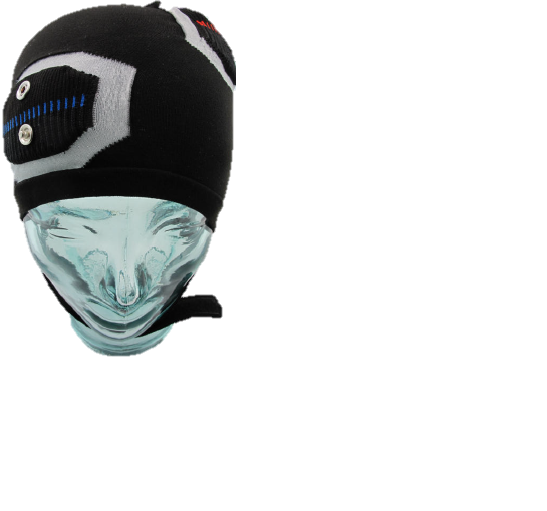 |
| 5) When the **cap** fits properly, close the **velcro under** the chin. | 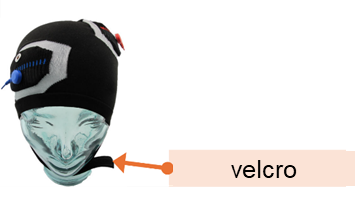 |
| 6) Now slowly fill **saline solution** with a **syringe** into the openings next to the electrodes. Use 12 ml of saline solution per electrode. The electrodes should be completely moistened, but the saline solution should not run outwards. | 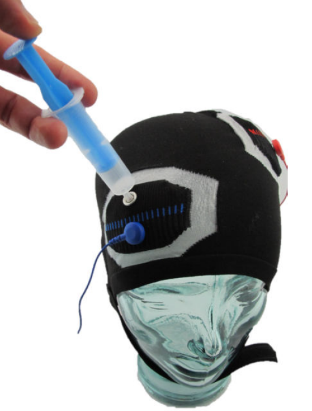 |

**Important:** The hood must not be moistened outside the electrodes.

| 7) Attach the **electrode cables** to the corresponding push buttons on the **cap**. Be sure to pay attention to **the coloured marking** (= red cable to red cap marking, blue cable to blue cap marking). | 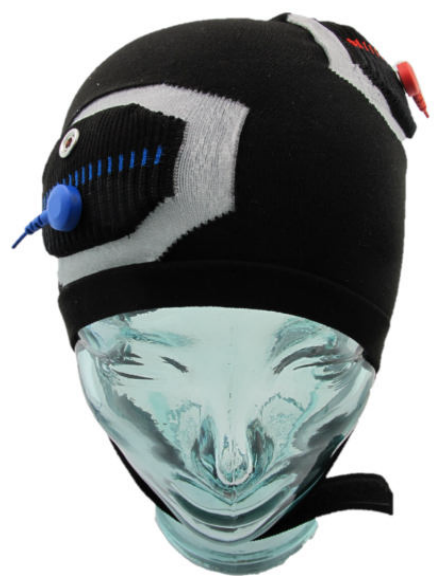 |
| --- | --- |
| 8) Connect the **stimulator** to the **electrode cables.**   Pay attention to the coloured marking (= red cable in red marked opening, blue cable in blue marked opening). Hold the cables by the connector, not the thin part of the cable. | 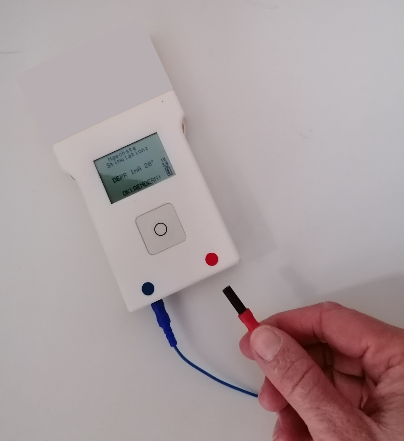 |
| 9) Check the mirror to make sure everything is connected correctly. Check the following points: a) Is the **cap** pulled up to the eyebrows? b) Are the **cables** placed in the correct color? (red = your left side of your body, blue = your right side of your body) c) Have both **electrodes** been moistened **with the** saline solution? d) Are **electrode cables** connected to the **cap and** to the **stimulator**? | |

If you can answer "yes" to all the questions, you can move on to the next step (logging into teletherapy).

### 4. Stimulation and therapy

#### 4.1 **Logging into teletherapy**

Required items:

- Stimulator (connected to the cap via the electrode cables)
- Memory module
- Cap (put on and connected to the stimulator via the electrode cables)
- Laptop

| 1) Sit at your desk with the **cap** on and the **stimulation device** and turn on the **laptop**. The power button is located on the left above the keyboard. | 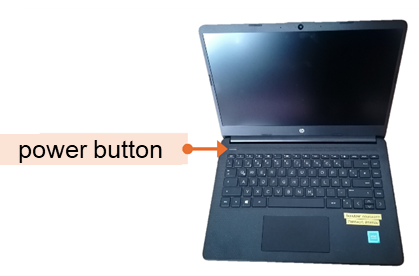 | |
| --- | --- | --- |
| 2) Enter the **username** and **password**. Both information can be found on the right below the keyboard. | 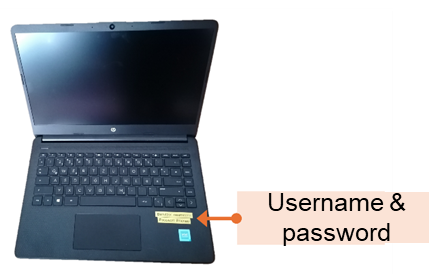 | |
| 3) Now go to your e-mail inbox. Open the **e-mail** of the study speech therapist. Click on the link. The link will take you to the BigBlueButton entry page. | 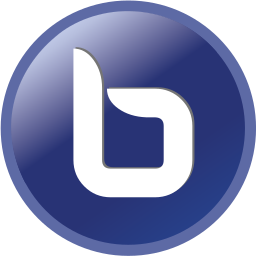 | |
| 4) Enter your **name** and **password** here. You will also find the password in the e-mail. | 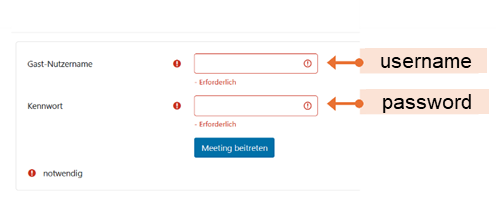 | |
| 5) Wait briefly in the **waiting area** of BigBlueButton until your speech therapist starts the therapy. | 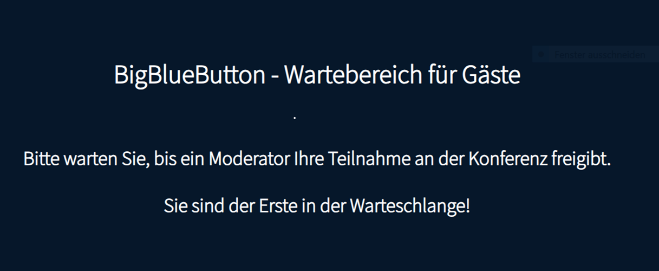 | |
| 6) Please allow BigBlueButton to access your **microphone** and **camera** by clicking "Allow". | 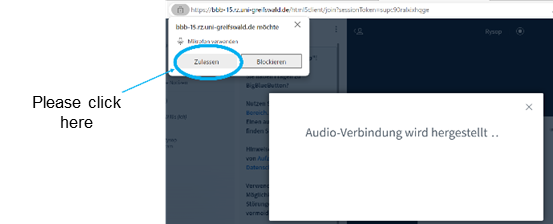 | |
| 7) Talk to the speech therapist to test whether the camera and microphone are working. | 8) Your therapist checks via the video whether the cap has been put on and connected correctly. | |
| 9) Now turn on the **stimulator** by connecting the **memory module** to the **stimulator** .  Make sure that the unlabeled pages face you. The stimulator turns on automatically. | 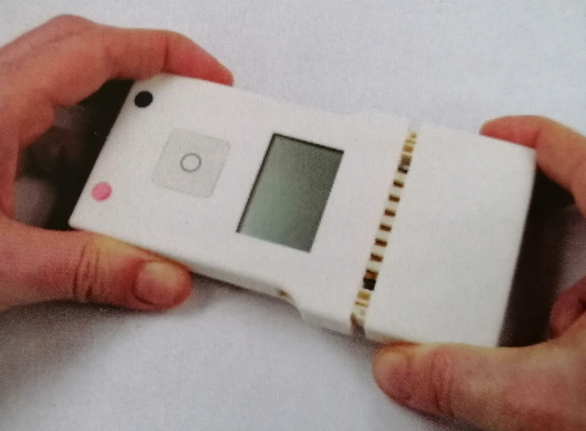 | |
| 10) On the **screen** of the **stimulator**, you will now see information about the stimulation. To confirm, briefly press the **button** on the stimulator 1 time. | 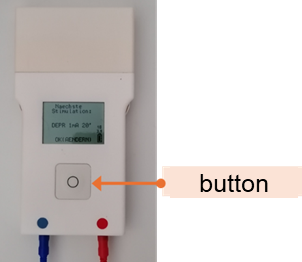 | |
| 11) Before stimulation begins, it is automatically checked whether **current flow** is possible (= **impedance control**).  The text on the screen will inform you of the result of the impedance check.  If the impedance value is OK, "OK", "START" will appear on the screen.  If the impedance value is not correct, an error message will appear (e.g. "HIGH IMPEDANCE"). Please read Chapter *7 Frequently Asked Questions to find out* what you need to do if you make a mistake. | | 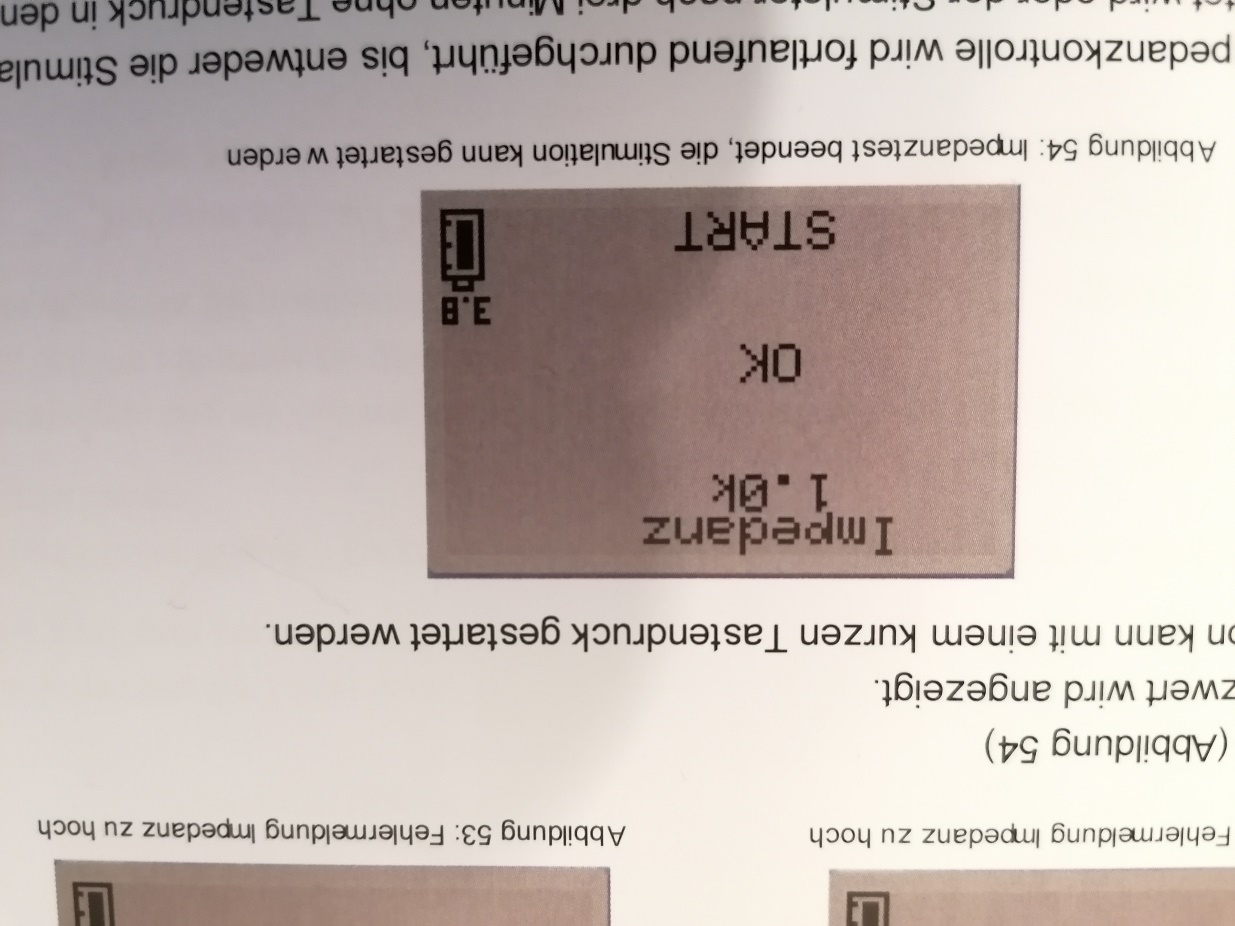 |

**Now all preparations are complete, and the therapy can begin.**

#### 4.2 Start therapy and stimulation

| 1) Your study speech therapist will let you know when you can **start the stimulation**. You can start the stimulation by **pressing** the button on **the stimulator**. | 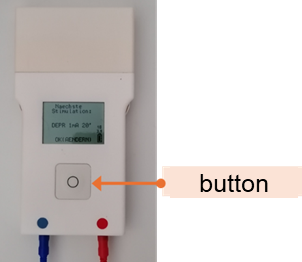 |
| --- | --- |
| 2) The **stimulation** ends automatically after **20 minutes**. The device switches off automatically.  The **therapy** goes even further. You can keep the cap on. | 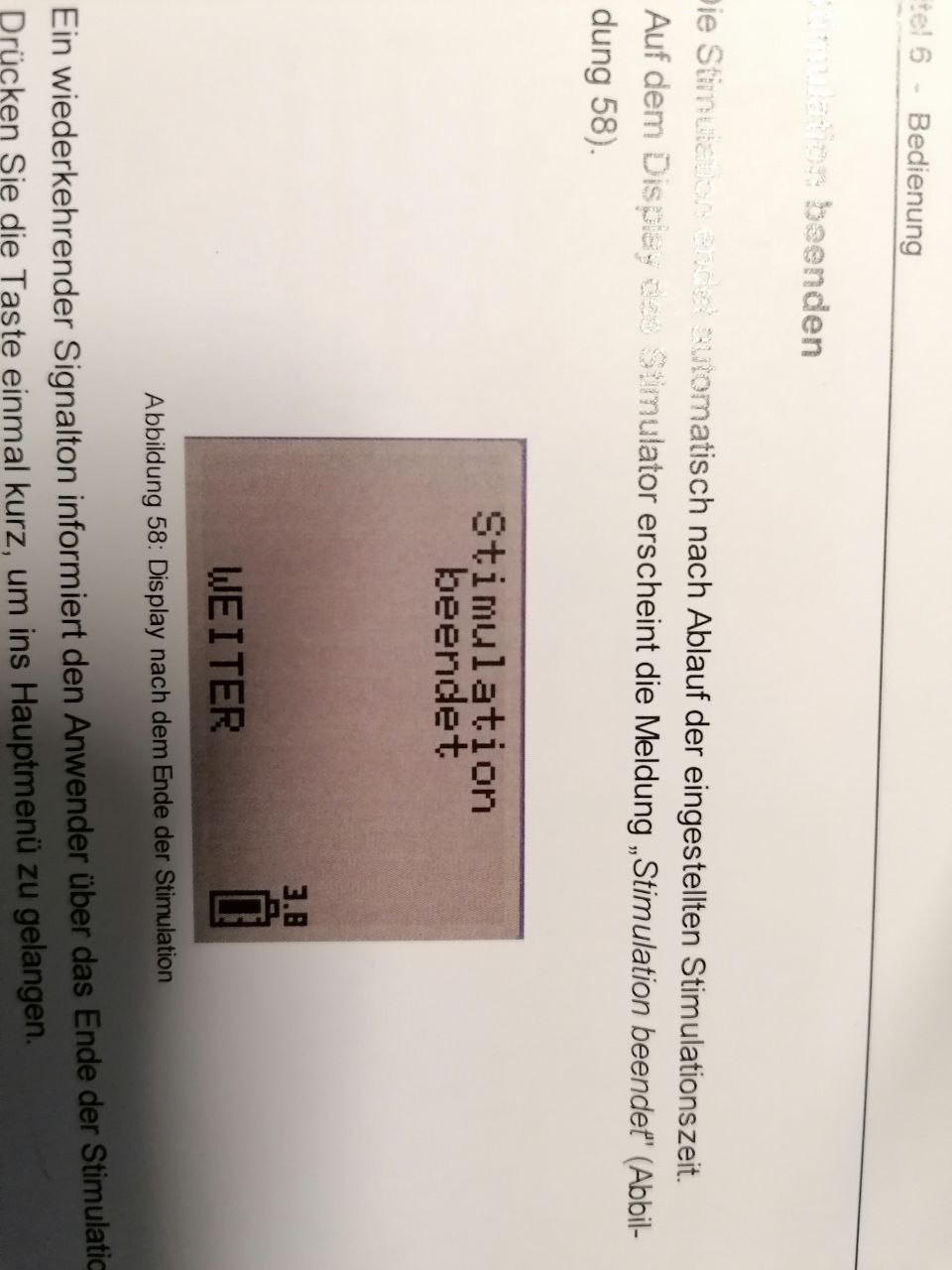 |

If you need a break during therapy, please let the study speech therapist know.

### 5. Follow-up of stimulation

After completion of the therapy and stimulation, the equipment is **dismantled**, **cleaned** and **stored** .

| 1) Remove the **cap** . To do this, open the **Velcro fastener** on the chin and carefully pull the cap upwards from your head. | |
| --- | --- |
| 2) Carefully remove the **push-button cables** from the electrodes on the cap. | |
| 3) Unplug the **cables** from the **stimulator**. To do this, please touch the socket at the end of the cable. | 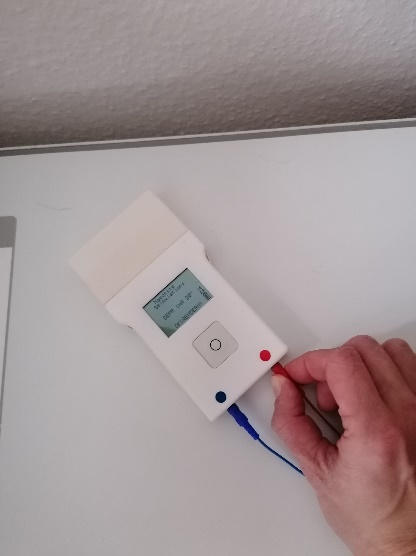 |
| 4) Remove the **memory module** from **the stimulator**. To do this, squeeze the two side locking springs together and at the same time carefully pull out the memory module. | 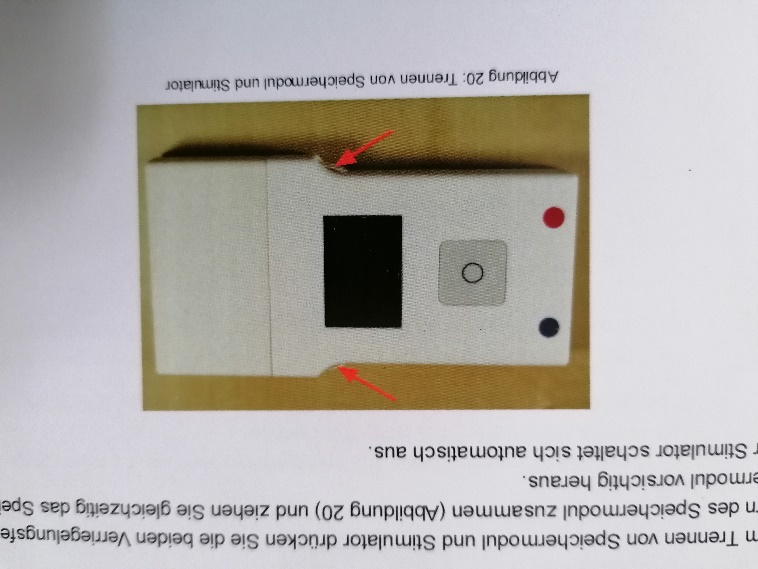 |
| 5) Connect the **memory module** to the **charger** and charge it (see *Chapter 3.1 Charging the stimulator*). | 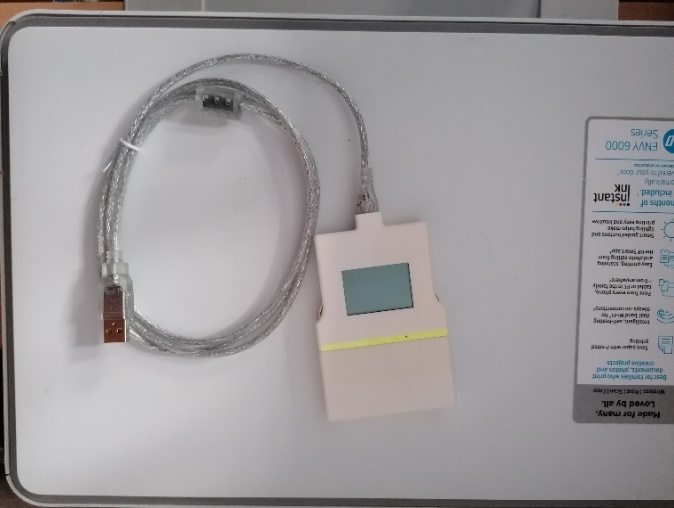 |
| 6) Wash the **cap** gently by hand with wool detergent or hygiene detergent. | |
| 7) Hang the **cap** to dry. For example, you can hang the cap over a bottle and place it near a heater. | |

**Important:** Only air dry the cap. Do not use a dryer or hair dryer under any circumstances and do not place the hood directly on a heater.

### 6. Big Blue Button

The following chapter gives you an overview of the most important functions of Big Blue Button, as well as hints and tips on how to use Big Blue Button.

Figure 3 shows the **main window** of Big Blue Button. This window appears when you have successfully dialed into the video conference. On the left, you can see the participants. In the middle you can see a large dark area, the **therapy area**. The image of your therapist appears on this surface and also your image if you have switched on the camera. Therapy content is also shown in this area.

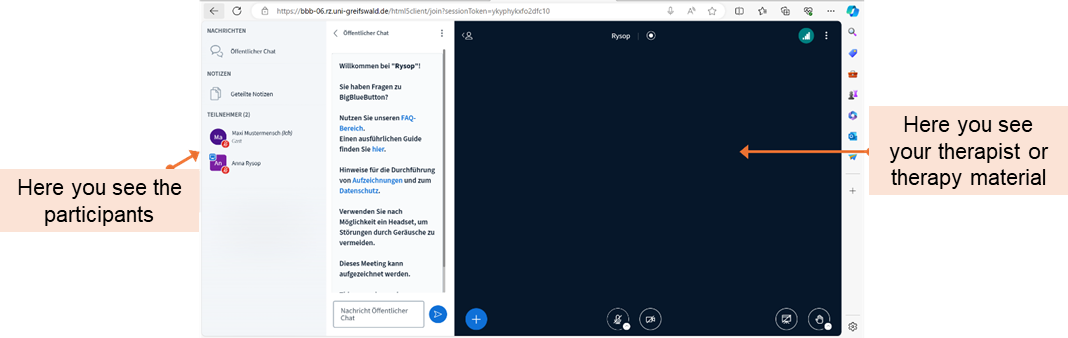

Figure 3. Big Blue Button main window.

Next, let us take a closer look at **the therapy area** (Figure 4). In the main window, you have various buttons that you can press. For example, you can turn your **microphone** and **camera** on and off. You can check how good or bad your **internet connection** is. But you can also turn the **whiteboard** on and off.

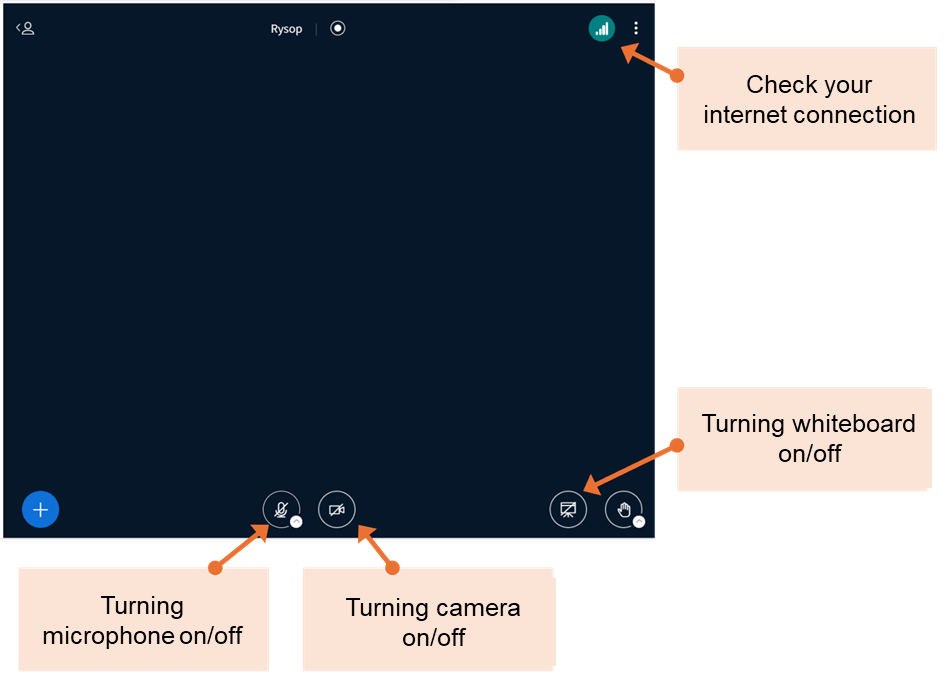

Figure 4: Therapy area of Big Blue Button.

You can think of the **whiteboard** as a **blackboard**. You can use it to write or draw **together with your therapist**. Figure 5 shows you an overview of the whiteboard and the most important functions of the whiteboard.

You have several **tools** to choose from. The **pen tool** allows you to **write** or **draw** freehand. To do this, select the pen tool with a mouse click and move the mouse pointer to the white area. Hold down the mouse pointer and move the mouse to draw. You can also write in this way. You also have another tool for writing, the **text tool**. To do this, select the Type tool (denoted by T) and click anywhere on the whiteboard. A text field will now open here, in which you can enter a text using the keyboard. To make the text larger, drag the edge of the text box.

If you want to delete something written or drawn, click on the **Eraser** tool and then on the object you want to delete.

But you can also simply move something written or drawn. To do this, press the **mouse cursor** tool and then click on the object you want to move.

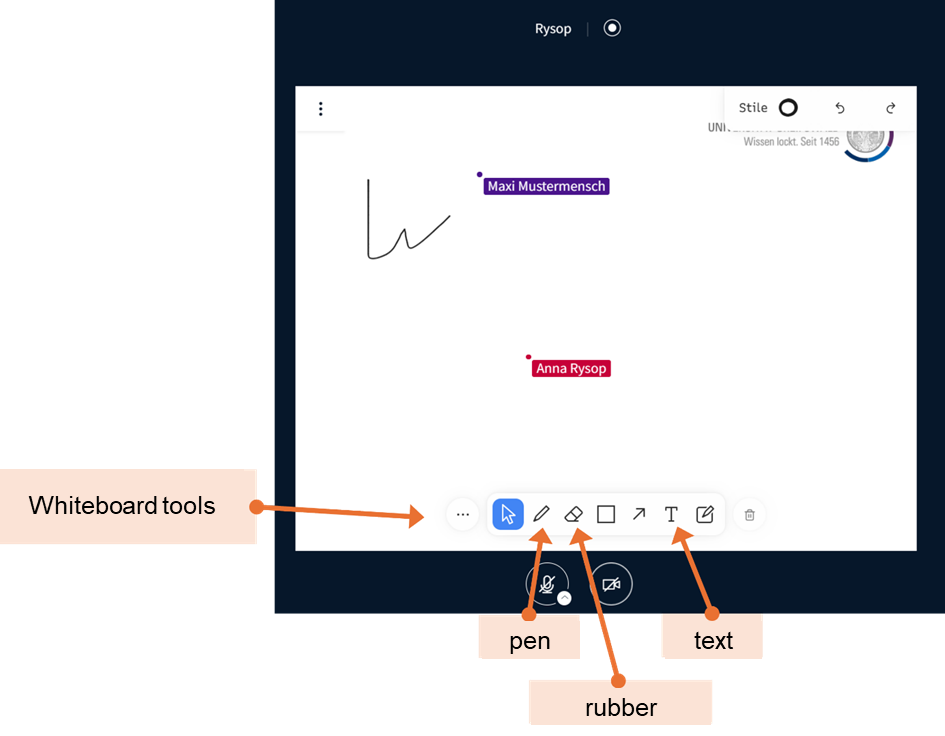

Figure 5: Whiteboard and tools of whiteboard.

### 7. Frequently Asked Questions

The following is a list of frequently asked questions and solutions to any questions and problems that may arise. If you have any further questions or problems, you can reach us at any time via the study e-mail address.

**Devices**

- How do I distinguish a charger and a programmer?
  - The charger has a yellow mark on the case.
- Laptop won't turn on
  - The battery may be empty. Charge the laptop using the included power adapter.
- Laptop stops responding
  - Shut down the laptop by pressing the power button for a longer period. Then start the laptop again.

**Stimulation**

- What error messages can occur during impedance control and how can they be resolved?
  - **"Impedance ??? – Connect electrodes**: This error message means that the electrode cables are not connected properly. Check that the cables are properly connected to the stimulator and cap.
  - **Impedance 49K - High impedance**: This error message means that the impedance is too high. If necessary, brush aside any hair directly under the electrodes. Check the impedance again. If this is still too high, carefully add some saline solution.
- Can the stimulation be interrupted?
  - The stimulation cannot be paused. However, it can be interrupted by pressing the button on the stimulator during stimulation.

**Teletherapy**

- Where can I find the link for teletherapy?
  - You will find the link in your e-mail inbox.
- What should I do if the sound and/or image do not work?
  - Please close the BigBlueButton window again. Click on the link in your emails again.
  - If the sound and/or picture still do not work, please restart the laptop.
